# TALQ: A reproducible language model questionnaire for dense measurement of depressive and anxious states

**DOI:** 10.64898/2026.11.12.26367797

**Authors:** Maxine E. Levesque, Kira E. Poskanzer, Joshua Woolley, Reza Abbasi-Asl

## Abstract

Depression and anxiety are often assessed weeks or months apart with brief questionnaires covering few symptoms, leaving changes between assessments unobserved. Here, we introduce TALQ, a scoring system in which a large language model rates 52 symptoms from text, and use it to score 491 members of two online mental health communities every two weeks for a year. Ratings were stable across samples, predicted human annotations of interpersonal risk factors, low self-esteem and depression severity, and agreed with inventories completed by the same model. The scores mapped into 12 replicable symptom factors, including fearful preoccupation and social strain. Sleep disturbance and self-harm content consistently preceded overwhelm and emotional pain, respectively, two weeks later. People differed mainly in the states they occupied rather than transitions between them, and 13% entered in a crisis that remitted over months. Overall, our findings suggest that TALQ can measure expressed mental states densely and quantify longitudinal dynamics.

---

Depression and anxiety disorders are among the largest causes of disability worldwide.^1^ Antidepressants are modestly more effective than placebo,^2^ and fewer than half of patients remit after a first course of treatment.^3^ These limited outcomes have been traced to how depression is defined and measured.^4^ Hundreds of symptom combinations satisfy the same diagnostic criteria,^5^ common rating scales have little content overlap,^6^ and sum scores obscure effects of treatment on specific symptoms.^7^ Symptoms are also measured only at visits weeks or months apart, if at all,^8^ so most of what happens between assessments goes unobserved. In network and dynamical accounts, symptoms influence one another over time, and the longitudinal dynamics of mental states may themselves characterize a disorder.^9–12^ Studying these dynamics requires repeated reliable measurement of many symptoms in many people over long periods.^13^ Clinical trials rely on clinician-administered scales such as the Montgomery-Åsberg Depression Rating Scale (MADRS)^14^, and routine care on questionnaires such as the Patient Health Questionnaire (PHQ-9)^15^ and the Beck Depression Inventory (BDI-II),^16^ but both cover few symptoms at visits usually weeks or months apart. Experience sampling provides dense data within individuals, but usually for a few items over days or weeks.^11^ Passive sensing with smartphones records behavior continuously, but mood and other experiences have to be inferred from it.^17,18^

People describe their mental states in their own words, for example in online communities, and the language of social media carries clinically relevant signal.^19–21^ Large language models (LLMs) annotate text with an accuracy approaching that of human annotators on many tasks^22,23^ and can infer mental health conditions from text without training on the task.^24,25^ Unlike a human rater, an LLM can be fixed and versioned, applied again to the same text, given any questionnaire and prompted repeatedly to quantify its own uncertainty.^26,27^ Dense ratings of naturalistic text also carry risks. People post irregularly, so analyses that assume evenly spaced observations can misjudge how symptoms change over time. It has not been established whether LLM ratings of such text are reliable and valid, what structure they have or whether they support claims about symptom dynamics.

Here we designed a mental health scoring system in which an LLM acts as a structured rater that answers a symptom questionnaire about the author of a set of posts, which we call TALQ (text assessment with language model questionnaires). We rated 52 symptoms compiled from depression rating scales^6^ every two weeks during the first year of sustained activity of 491 members of two Reddit communities, r/depression and r/Anxiety. We assessed the reliability and validity of the ratings, identified 12 replicable mental health factors and examined how they change within each person, across weeks and months.

## Results

### TALQ, a mental health scoring system built on an LLM

Scoring mental states from what people write requires a rater that applies the same symptom items in the same way to every person and period. We therefore built TALQ, a scoring system in which an LLM rates the author of a text on 52 symptoms compiled from seven depression rating scales,^6^ namely the MADRS, BDI-II, Hamilton Rating Scale for Depression (HRSD)^28^, Center for Epidemiologic Studies Depression Scale (CES-D)^29^, Inventory of Depressive Symptomatology (IDS)^30^, Quick Inventory of Depressive Symptomatology (QIDS)^31^ and Zung Self-Rating Depression Scale (SDS)^32^ (**Fig. 1a**). The 52 symptoms come from a content analysis of the combined 125 items of these scales that merged items with the same content.^6^ For each of these 52 predefined symptoms, the LLM reads all text a person wrote during a period and returns a rating from 0 (absent) to 100 (present), a short justification and its confidence for each symptom (**Supplementary Note 1**). Within each period, the LLM rates each symptom several times from the same text, and these repeated ratings are averaged into one score for that period. Here we applied TALQ to posts and comments by 491 users of two online communities, grouped into windows of two weeks and rated by Llama-3.1-8B-Instruct, an LLM with 8 billion parameters.^33^ The users had 9,886 two-week windows with any posts or comments (median 22 per user and 417 words per window, **Extended Data Fig. 1** and **Methods**), and rating each window on all 52 symptoms five times yielded 2.54 million valid ratings (**Fig. 1b** and **Extended Data Fig. 2**).

**Fig. 1.**
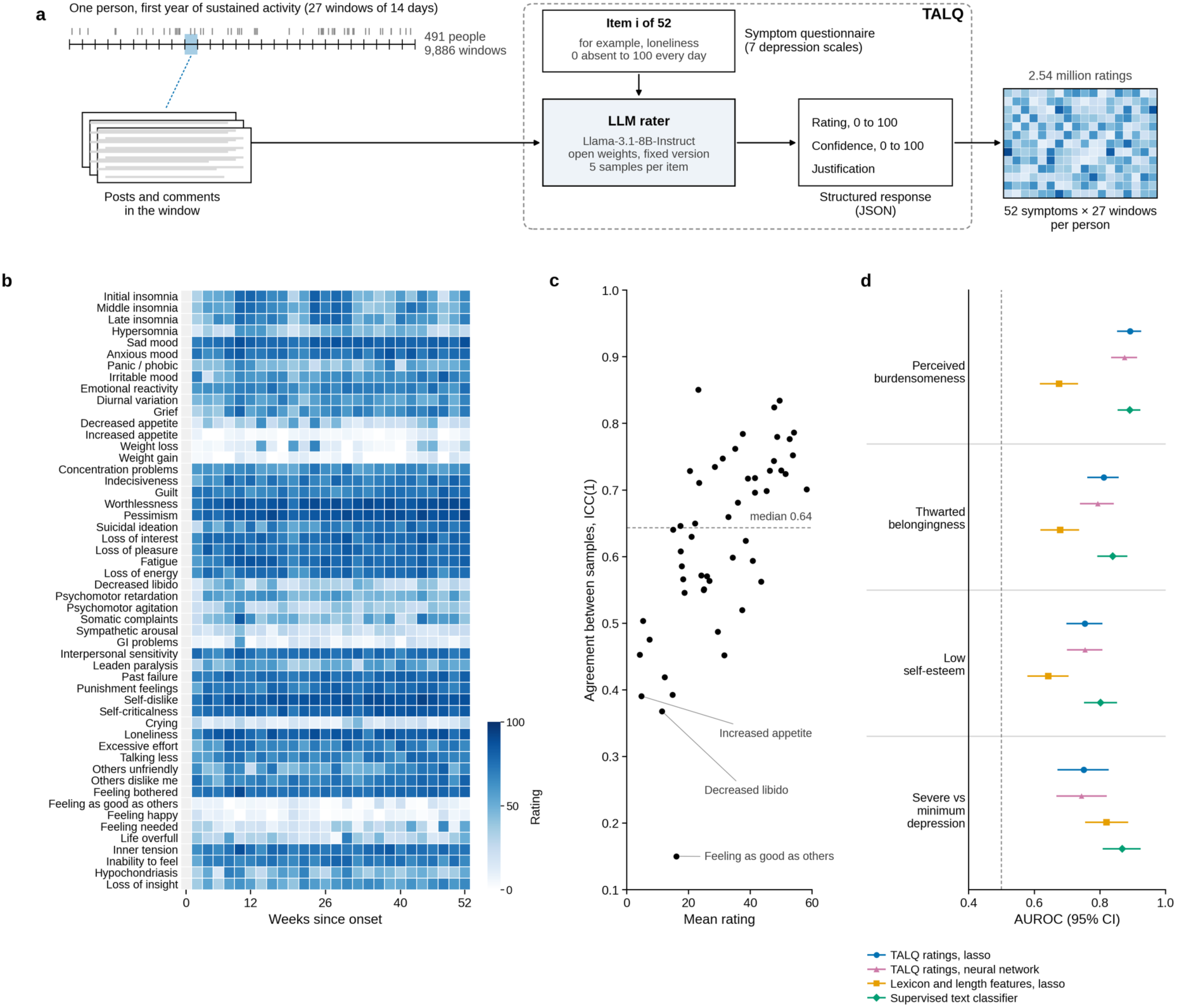
TALQ, a mental health scoring system built on an LLM. **a**, Design, with the components of TALQ in the dashed frame. Users with at least 360 active days in r/depression or r/Anxiety were followed through 27 windows of two weeks from the onset of sustained activity. For each window, the LLM rater (Llama-3.1-8B-Instruct) received all posts and comments of the person and one of 52 symptoms compiled from depression rating scales,^6^ and it returned a rating from 0 to 100, its confidence and a short justification, five times per window and symptom. **b**, Ratings (mean of five samples) of all 52 symptoms across the year of one illustrative user, in the order of **Supplementary Table 1**, where the full titles of the abbreviated names are given. Light gray cells mark windows without posts. **c**, Agreement between the five samples of the rater, shown as the ICC(1) of single ratings for each symptom against the mean rating of that symptom. The dashed line marks the median. **d**, Prediction of human annotations for four labels in three datasets of single Reddit posts, scored with the same prompt by a local deployment of the rater. The ratings of all scored symptoms of a post, each the mean of five samples, are combined by a lasso (blue) or by a neural network with one hidden layer (pink), cross-validated over posts, and compared with nine lexicon and length features combined by the same lasso (orange) and with a supervised classifier (green), a logistic regression on term frequency–inverse document frequency features of the words of the posts, trained on the labels with fivefold cross-validation. Points show the area under the receiver operating characteristic curve (AUROC), lines show 95% bootstrap intervals and thin rules separate the four labels. For severity, the AUROC compares severe with minimum depression.

To obtain a reliable and robust^34^ score for each symptom, we prompted the LLM five times with the same input for each window and symptom and took the average of the five ratings per symptom. The five ratings differed only by the randomness with which the model generates its answers. We found that across the five repeats, the ratings agreed moderately (median intraclass correlation, ICC, 0.64 across symptoms, range 0.15–0.85, **Fig. 1c**), but the mean of all five was highly reliable (median ICC of the mean 0.90). Therefore, we used the mean as the score of each window and symptom. Because the posts of a window may not contain enough information to rate every symptom, we also used the confidence that the model reported with each rating. Our main analyses used the 7,094 windows from 481 users in which the mean confidence exceeded 40 of 100 for all 52 symptoms (**Extended Data Fig. 1**). We chose this threshold by examining the LLM’s generated confidence scores, which tracked the reliability of its ratings across five repeats (**Supplementary Table 4**). Agreement between the five ratings of a window and symptom rose steadily with confidence, from a median ICC of 0.19 for confidence scores below 40 (0.5% of ratings) to 0.90 for those above 80. We chose a threshold of 40 to exclude windows with the least reliable ratings while keeping 73% of complete windows.

The users in our cohort are anonymous, so no clinical assessments are available for them, and no public dataset has human annotations of all 52 symptoms. We therefore asked whether TALQ ratings can predict available human annotations of single posts in three other Reddit datasets,^35–37^ where we have access to the following annotations: Two interpersonal risk factors, (1) thwarted belongingness and (2) perceived burdensomeness, as well as (3) low self-esteem and (4) depression severity. As in the cohort, each post was rated five times on each of the symptoms associated with the annotations (see **Methods**), and the five ratings were averaged. We used a regularized regression model (lasso)^38^, cross-validated over posts, to predict the human annotations from these ratings. The model discriminated perceived burdensomeness (area under the receiver operating characteristic curve, AUROC, 0.89, 95% CI 0.85–0.93), thwarted belongingness (AUROC 0.81, 95% CI 0.76–0.86) and low self-esteem (AUROC 0.75, 95% CI 0.70–0.81), and separated severe from minimum depression (AUROC 0.75, 95% CI 0.67–0.83) (**Fig. 1d**). In a symptom importance analysis, we counted a symptom as important when the lasso kept it in at least 80% of the 50 training folds (**Extended Data Fig. 3a,b**). Perceived burdensomeness relied on suicidal ideation alone, and the other labels mainly on loneliness (thwarted belongingness), worthlessness and self-dislike (low self-esteem) and concentration problems, psychomotor agitation and suicidal ideation (depression severity). The first three annotations (perceived burdensomeness, thwarted belongingness and low self-esteem) each corresponded to one symptom, which alone predicted the annotation label as well as all symptoms combined (suicidal ideation AUROC 0.89, loneliness 0.80 and self-dislike 0.73, against 0.89, 0.81 and 0.75 for the lasso). Depression severity drew on eight symptoms, but combining them did not significantly improve on the best one, concentration problems (AUROC 0.75 against 0.74, difference 0.01, 95% CI −0.04 to 0.05). To test whether a nonlinear model could improve the predictive performance, we also trained a small neural network (multilayer perceptron), which did not improve on the lasso (AUROC 0.88, 0.79, 0.75 and 0.74, within 0.02 of the lasso, **Fig. 1d**).

As a baseline that needs no LLM, we used nine lexicon and length features, which score each post with published sentiment and emotion word lists (lexicons), count first person singular pronouns and absolutist words such as “always” and measure post length (**Methods**). Using lasso, they predicted perceived burdensomeness, thwarted belongingness and low self-esteem less well than the TALQ ratings (AUROC 0.64–0.68 against 0.75–0.89, differences 0.11–0.22, each 95% CI above zero) but depression severity at least as well (AUROC 0.82 against 0.75).

To validate TALQ against the choice of LLM, we scored the same posts once with Qwen3.6-35B-A3B,^39^ a larger model with open weights and 35 billion parameters. This LLM never declined to answer, whereas Llama-3.1-8B declined 1.1% of requests because of its safeguards. Compared with a single Llama-3.1-8B-Instruct rating (the first of its five), the new ratings predicted every annotated label at least as well. The prediction was better for low self-esteem (AUROC 0.80 versus 0.72, difference 0.08, 95% CI 0.04–0.13, **Extended Data Fig. 3e**). Because clinical care and research rely on standardized self-report inventories, we also had the same LLM (Llama-3.1-8B-Instruct) complete the BDI-II and PHQ-9 for each window as a proxy respondent. In this case, the LLM answered each item in the first person on behalf of the author of the posts. To compare these inventory ratings with TALQ symptom scores, we built a TALQ analog of each inventory. Before any inventory rating was analyzed, we matched each inventory item to the TALQ symptoms with the same content (**Supplementary Table 3**). For example, we matched the BDI-II item on sadness to sad mood and the PHQ-9 item on anhedonia to loss of interest and loss of pleasure. The TALQ analogs agreed closely with the inventory totals (ρ = 0.90 and 0.92 across 1,672 and 1,650 windows, and 0.82 and 0.85 within users, **Extended Data Fig. 4**). Because the same LLM produced both the TALQ ratings and the inventory answers, this agreement shows consistency across questionnaire formats and not necessarily the agreement with an independent rater.

### Twelve replicable mental health factors

To identify the main dimensions along which the 52 TALQ symptoms vary together, we asked whether a small number of latent factors could explain the symptoms. To answer this question, we used nonnegative matrix factorization (NMF) to decompose the ratings into K = 12 factors. Among K = 11 to 14, K = 12 was the most stable across random initializations by the staNMF criterion^40,41^ (instability 0.015, against 0.058, 0.034 and 0.045 for K = 11, 13 and 14, **Fig. 2b**). Solutions with 10 or fewer factors or with 18 were also stable but merged physical distress with anhedonia (K = 10) or split off factors dominated by single symptoms (K = 18, **Extended Data Fig. 5e**). The 12 factors (**Fig. 2a** and **Table 1**) were Anxious tension, Fearful preoccupation, Social strain, Positive wellbeing, Sleep disturbance, Physical distress, Emotional instability, Overwhelm, Anhedonia, Emotional pain, Negative self-reflection and Self-harm tendency. Together they covered the nine criterion A symptoms of major depressive disorder and the core symptoms of generalized anxiety disorder in the Diagnostic and Statistical Manual of Mental Disorders (DSM-5-TR).^42,43^ This was expected from a battery compiled from the seven depression rating scales. Interestingly, our data-driven factor analysis grouped these symptoms in ways the diagnostic criteria do not. Social strain, the sense of being judged negatively by others, had no diagnostic counterpart, and anxiety split into Anxious tension and Fearful preoccupation, which was dominated by illness anxiety, panic and sympathetic arousal. Domains are useful only if they recur in independent samples and are measured consistently, and the factors largely met both standards. Factors fitted separately in two random halves of the users matched each other closely (median Tucker congruence φ 0.98 over 20 splits, 12 of 12 factors above 0.85, **Fig. 2c**). Eleven factors were also internally consistent (weighted Cronbach’s α 0.93–0.98, **Fig. 2d** and **Table 1**), whereas Positive wellbeing was not (α 0.52). The structure differed more between the 370 users who wrote mostly in r/depression and the 111 who wrote mostly in r/Anxiety than between random groups of the same sizes (median φ 0.88 against 0.98, six of 12 factors below 0.85, **Extended Data Fig. 5b**). In the r/Anxiety group, where suicidal ideation was usually rated zero, Self-harm tendency had no counterpart (φ 0.14), and its place was taken by a factor of health anxiety that reshaped the other anxiety factors.

**Fig. 2.**
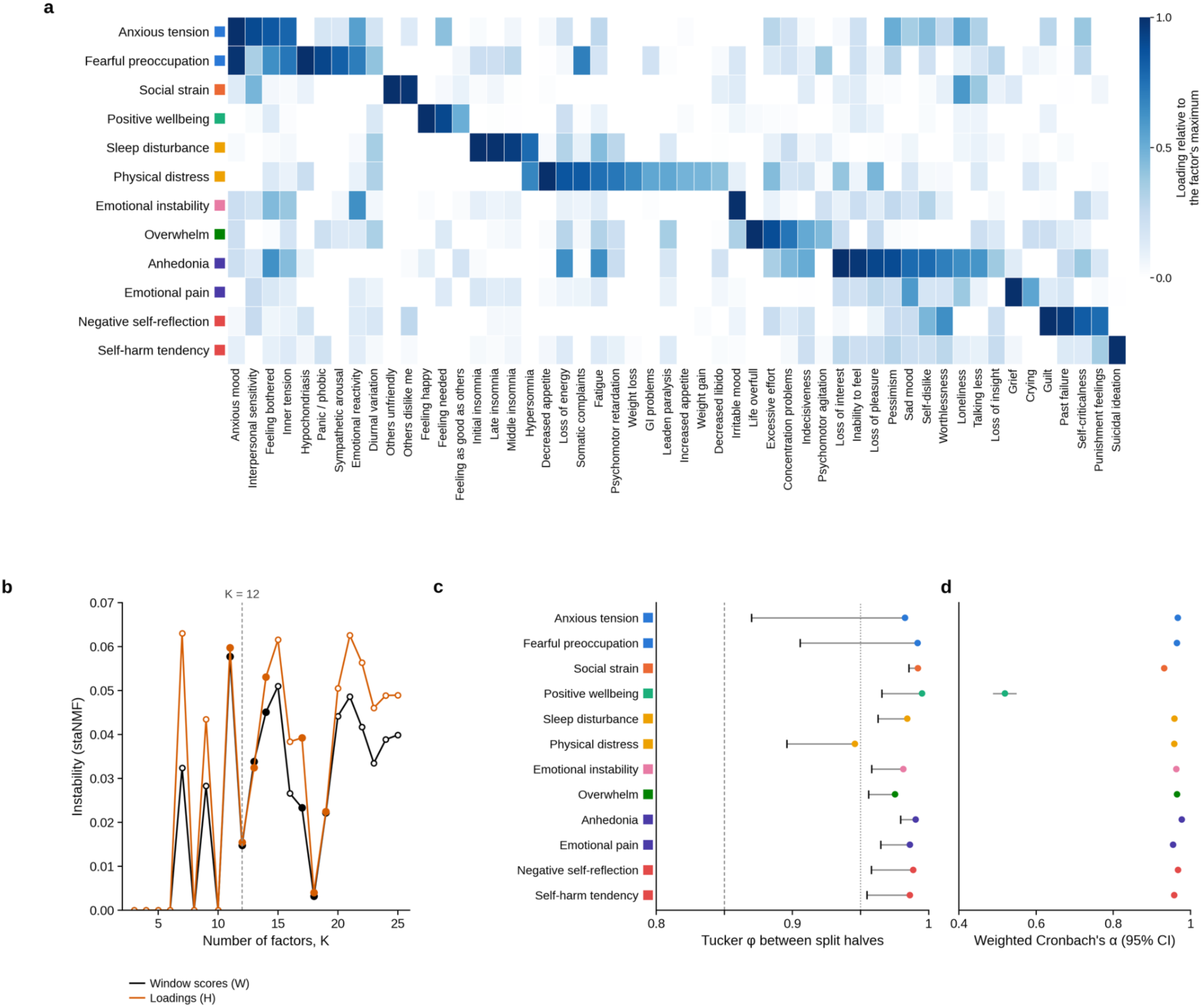
Twelve replicable mental health factors. **a**, Loadings of the 52 symptoms on the 12 factors of the nonnegative matrix factorization (K = 12), each row scaled to its maximum. Symptoms (columns) are grouped under the factor in which their relative loading is highest and are ordered within each factor from the highest down. Names are abbreviated as in Fig. 1b. Colored squares mark the clinical family of each factor (anxiety, social, positive affect, somatic, affective lability, cognitive load, depressive affect and self-referential), and the same colors identify the factors in the other figures. The share of each factor’s weight carried by its main symptoms is shown in **Extended Data** Fig. 5d. **b**, staNMF instability across numbers of factors, computed on window scores (W) and on loadings (H). Instability is the mean pairwise dissimilarity between solutions from different random initializations (100 initializations for filled markers and 20 for open markers). **c**, Replication across disjoint halves of the cohort, shown as the Tucker congruence of each factor between the loadings fitted separately in the two halves of 20 random splits of users. Dots give the median over splits and ticks give the 5th percentile. The dashed line marks 0.85 (fair similarity) and the dotted line 0.95 (equality).^44^ **d**, Internal consistency of each factor, the weighted Cronbach’s α of the composite weighted by the factor loadings (prefactor k_eff/(k_eff − 1)), with 95% intervals from 200 bootstrap resamples of users.

**Table 1.**
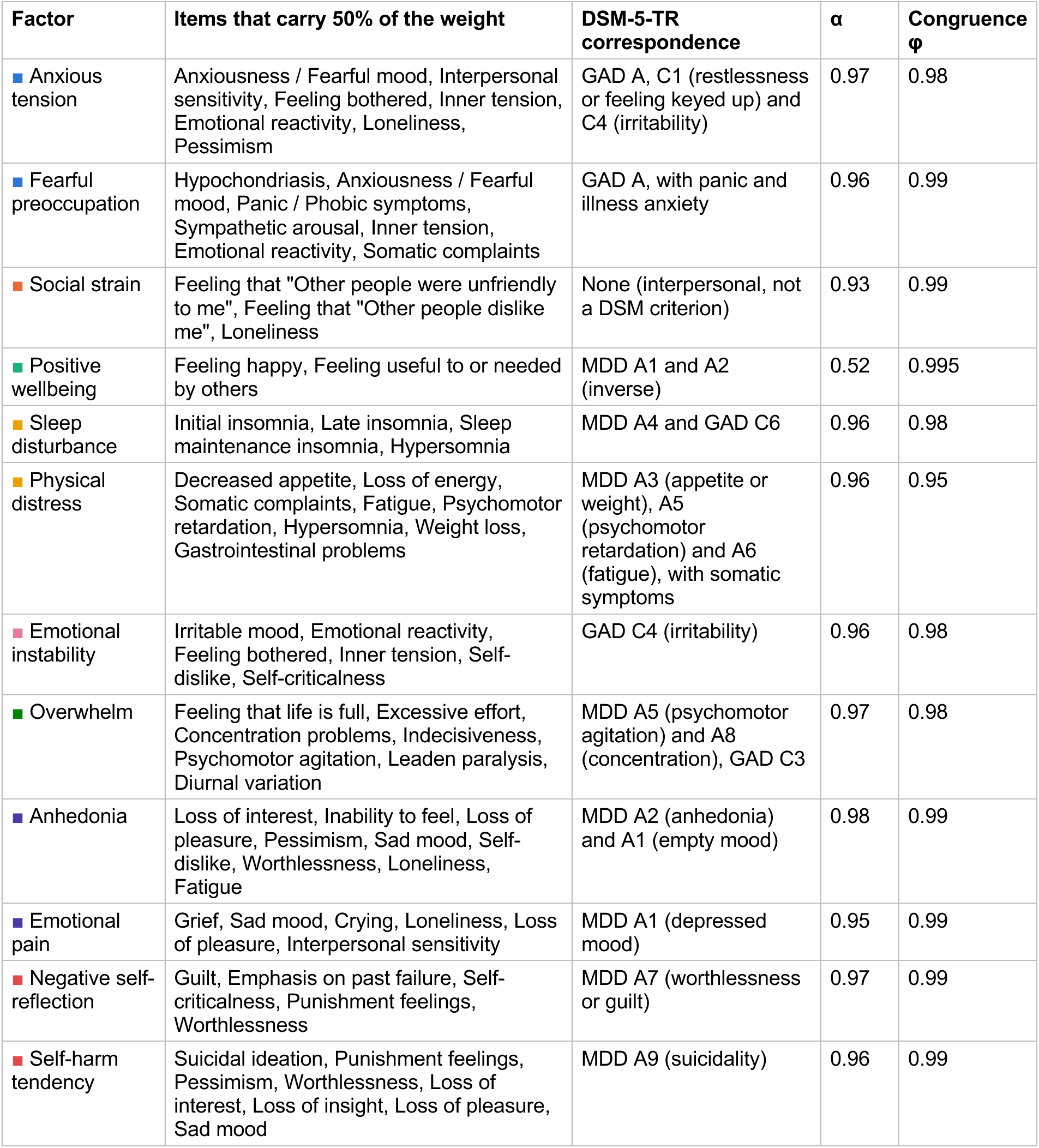
Mental health factors. For each factor, the table lists the items that together carry 50% of its NMF weight, the corresponding DSM-5-TR criteria for major depressive disorder (MDD) and generalized anxiety disorder (GAD),^42,43^ the weighted Cronbach’s α and the median Tucker congruence between split halves. Colored squares mark the clinical family of each factor as in **Fig. 2**. The authors chose the factor labels with assistance from the language model (**Methods**).

### Sleep disturbance and self-harm content precede overwhelm and emotional pain, respectively

Clinicians often ask which symptom comes first, for example whether poor sleep precedes low mood, because an early symptom can warn of relapse and may be a target for treatment. We tested whether one factor predicted others two weeks later with a vector autoregression (VAR) within users, corrected for the bias of short panels by the half-panel jackknife.^45^ Overall, little information carried over from one window to the next (autoregressive coefficients 0.02–0.15, **Fig. 3a**). However, of the 132 directed pairs of factors, 16 had P < 0.05, against 6.6 expected by chance, and two remained Granger predictive after control of the false discovery rate (FDR, **Fig. 3b** and **Extended Data Fig. 7a**). Both pairs had clear clinical content and ran in one direction. *Self-harm* content predicted more *emotional pain* two weeks later, whereas emotional pain did not predict later self-harm content (P = 0.54). This direction may seem counterintuitive, given theories that treat psychological pain as a driver of suicidal ideation,^46^ but it agrees with intensive longitudinal studies in which suicidal ideation predicted later hopelessness, negative affect and psychological pain over hours to days^47–49^ (see **Discussion**). *Sleep disturbance* likewise predicted more *overwhelm*, and not the reverse (P = 0.26). An impulse of 50 points raised Emotional pain by 4.5 points (95% CI 2.7–6.6) and Overwhelm by 3.9 points (95% CI 2.1–5.9), and both responses faded within six weeks (**Fig. 3c** and **Extended Data Fig. 7b**). Weaker pairs pointed the same way, with Sleep disturbance the earlier factor in 6 of the 16 pairs with P < 0.05 and Self-harm tendency in 4 (**Fig. 3b**).

**Fig. 3.**
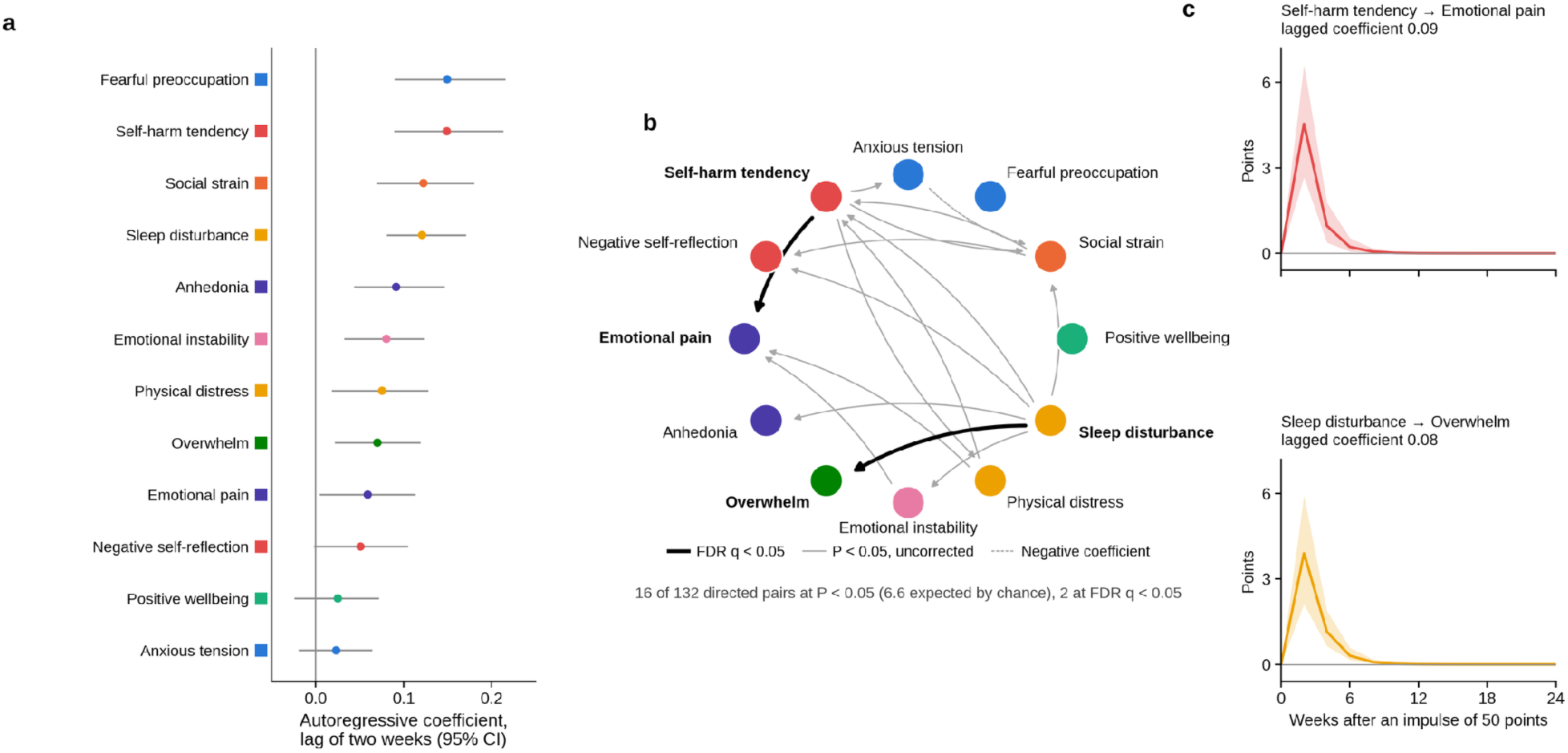
Sleep disturbance and self-harm content precede overwhelm and emotional pain. **a**, Autoregressive coefficient at lag 1 of each factor in a panel VAR(1) within users on exactly adjacent windows, corrected for the bias of short panels with the half-panel jackknife, with 95% intervals from a bootstrap over users. **b**, Lagged effects between factors. Arrows run from a factor in one window to the factor it predicted two weeks later. Black arrows mark the two pairs declared Granger predictive at a false discovery rate (FDR) of q < 0.05, and gray arrows the other pairs with uncorrected P < 0.05 (dashed for a negative coefficient), from Wald tests based on a covariance from a bootstrap over users. Bold names mark the factors of the two FDR pairs. **c**, Responses to an impulse of 50 points for the two pairs, with their lagged coefficients and 95% bands from a bootstrap over users. Colors identify the factors as in Fig. 2.

### Recurrent mental states

Clinicians often study the recurring states of people, such as a period dominated by panic or by hopelessness, and how long such states last informs how often a person should be seen. We therefore asked if we could identify such recurring states across symptom factors. We used a Dirichlet process Gaussian mixture model, with hyperparameters chosen by the Bayesian information criterion (BIC, **Extended Data Fig. 8**) and found 11 recognizable states (**Fig. 4a,b**). These included a common euthymic state of positive wellbeing with mild anxious tension (PW), a state combining self-harm content with social strain (SH+SS) and two states combining sleep disturbance with anhedonia (SD+AN). People stayed in a state hardly longer than expected from how often they occupied it (mean probability of remaining 0.25, against 0.23 with each user’s windows shuffled in time, P = 0.005, **Fig. 4c,d**).

**Fig. 4.**
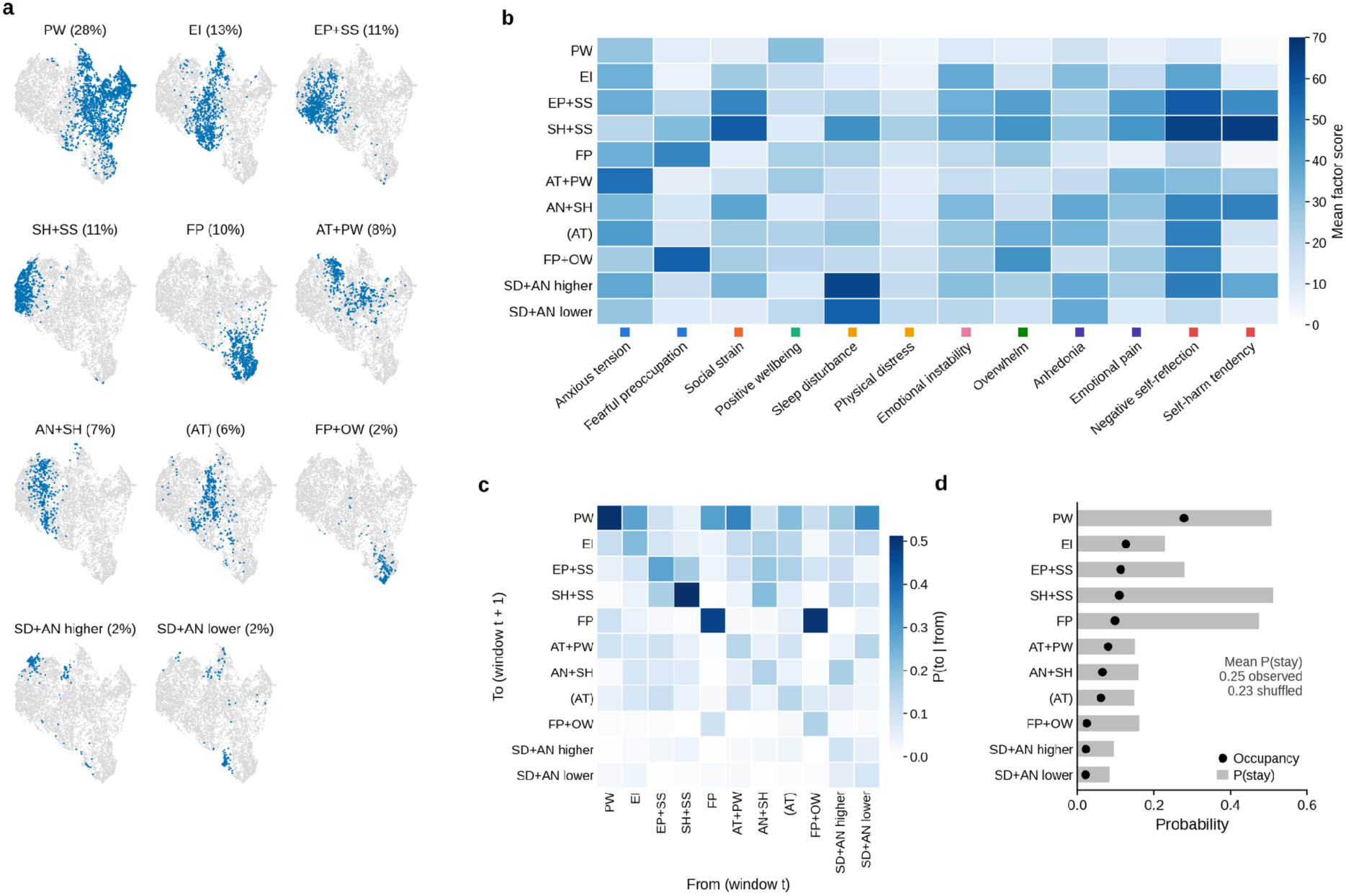
Recurrent mental states. **a**, Embedding of the factor profiles of all confident windows by uniform manifold approximation and projection (UMAP, trustworthiness 0.94). Each panel marks in blue the windows assigned to one state of the Dirichlet process Gaussian mixture, with the percentage of windows in parentheses. States are labeled by the factors at least one standard deviation above their mean across states, or by the highest factor in parentheses when no factor qualifies. The labels abbreviate the factors as AN (Anhedonia), AT (Anxious tension), EI (Emotional instability), EP (Emotional pain), FP (Fearful preoccupation), OW (Overwhelm), PW (Positive wellbeing), SD (Sleep disturbance), SH (Self-harm tendency) and SS (Social strain). When two states share a label, ‘higher’ and ‘lower’ mark the state with the higher and the lower mean of the factors other than Positive wellbeing. **b**, Mean factor scores of each state, with colored squares marking the clinical family of each factor as in Fig. 2. **c**, Population transition matrix between states in exactly adjacent windows, P(to | from). **d**, Probability of remaining in each state (bars) and occupancy of the state (dots). The inset compares the mean persistence with its value when each user’s windows are shuffled in time, which keeps the occupancy of every user (200 shuffles).

### Individuals differ mainly in the states they occupy over shorter time scales

If people differed in how they move between states, they could form dynamic subtypes with their own prognosis.^12^ We modeled each user’s sequence of states as drawn from one of several first-order Markov chains, or dynamical regimes.^50^ The Akaike information criterion corrected for small samples (AICc)^51^ selected four regimes, mostly euthymic, severe distress with social strain and self-harm content, fearful preoccupation alternating with euthymia, and mixed and mild (**Fig. 5d,e** and **Extended Data Fig. 9**). Mixtures that model occupancy alone predicted the sequences of users withheld from fitting as well as or better than Markov mixtures (**Fig. 5a**), and transitions added little beyond occupancy, although more than in sequences shuffled within each user (P = 0.005, **Fig. 5b**). A subtype is useful only if a person is assigned to it consistently, and regimes based on transitions were less reproducible than groups based on occupancy. When the two halves of each user’s year were classified by models fitted to other users, Markov regimes placed both halves in the same cluster for 50% of users (κ = 0.34, against 25% expected by chance) and occupancy mixtures for 67% (κ = 0.56, **Fig. 5c**). People were thus distinguished more reliably by the states they occupied than by their transitions.

**Fig. 5.**
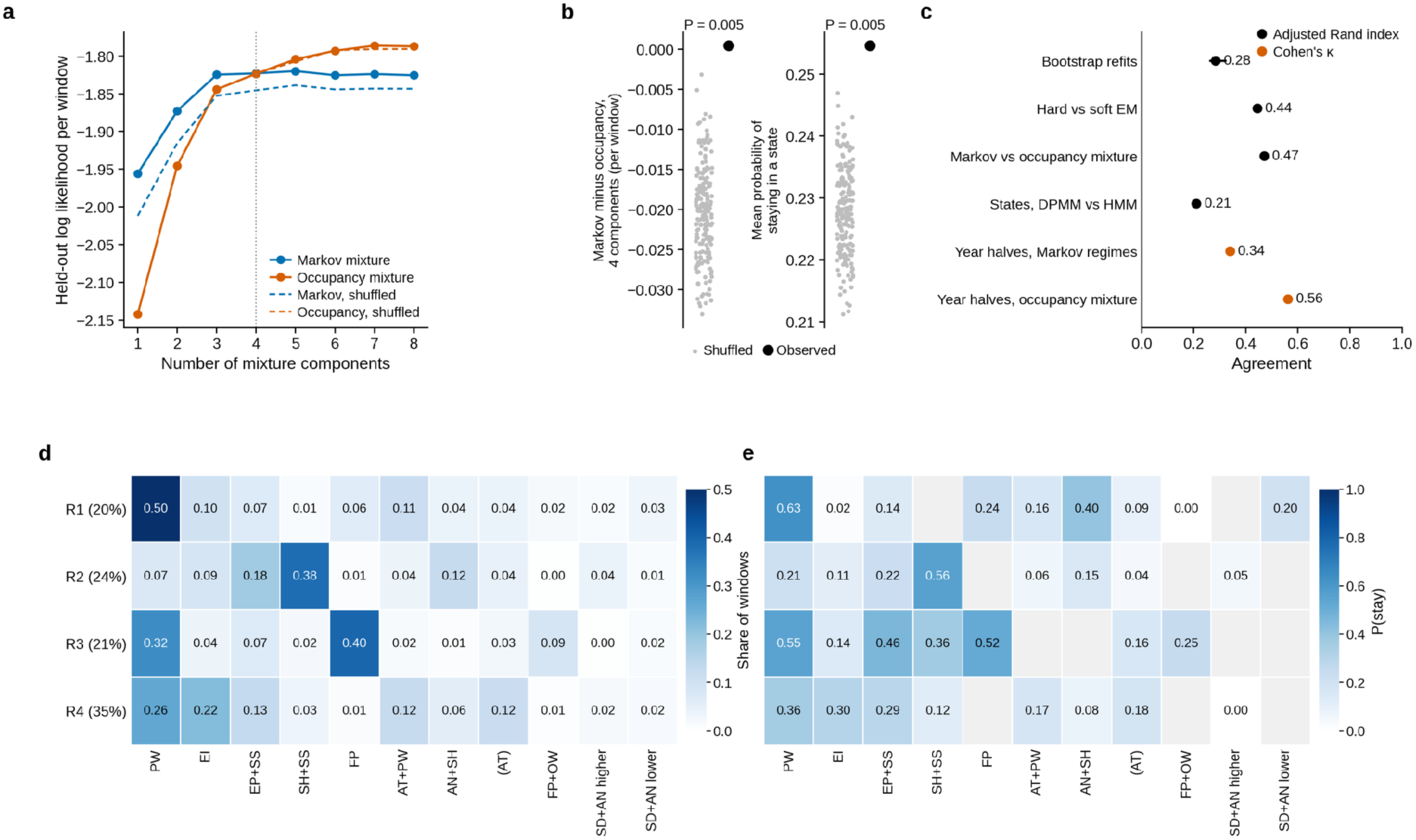
Individuals differ mainly in the states they occupy. **a**, Cross-validated log likelihood of the state sequences of users left out of fitting (five folds over users) for mixtures of first-order Markov chains (blue) and for mixtures that model state occupancy only (orange), by number of mixture components. Solid lines show the observed sequences and dashed lines the sequences after shuffling each user’s windows in time. The dotted line marks the number of regimes selected by AICc for Markov mixtures fitted with hard assignments (four). **b**, Difference in cross-validated log likelihood per window between Markov and occupancy mixtures at four components (left) and mean probability of staying in a state (right), observed (black) and in 200 shuffles within users (gray). **c**, Stability of the classifications. The adjusted Rand index (ARI) compares the main regime labels, from expectation–maximization (EM) with hard assignments, with labels from bootstrap refits (median and interquartile range), from EM with soft assignments and from an occupancy mixture. A further ARI compares the states of the Dirichlet process Gaussian mixture (DPMM) with those of a Gaussian hidden Markov model (HMM) fitted to the factor scores. Cohen’s κ compares the classifications of the first and second halves of each user’s year by Markov regimes and by occupancy mixtures, each fitted on the other users (244 users). **d**, Share of the windows of each regime that fall in each state. This occupancy also serves as the distribution of initial states of the regime, and row labels give the share of users. **e**, Probability of remaining in each state within each regime. Gray cells mark states with less than 2% of the windows of the regime.

### People differ in the course of their symptoms over months

Psychiatric classification and prognosis rest on the course of symptoms over months, such as whether an episode remits or persists. We therefore investigated the longitudinal course of the symptom factors over months. First, we examined overall severity, which we defined as the mean of the 11 factors other than Positive wellbeing. Overall severity was more alike in windows close in time than windows far apart (dissimilarity 0.79 and 1.16 times that of windows shuffled in time, at 2 and 38–44 weeks, **Fig. 6a**), suggesting change over longer time scales. The direction of each person’s change over the year was reproducible (r = 0.35 between slopes from the odd and from the even windows, against 0.00 after shuffling), whereas inertia and instability were not (r = −0.04 and 0.01 between the two halves of the year, **Fig. 6b**).

**Fig. 6.**
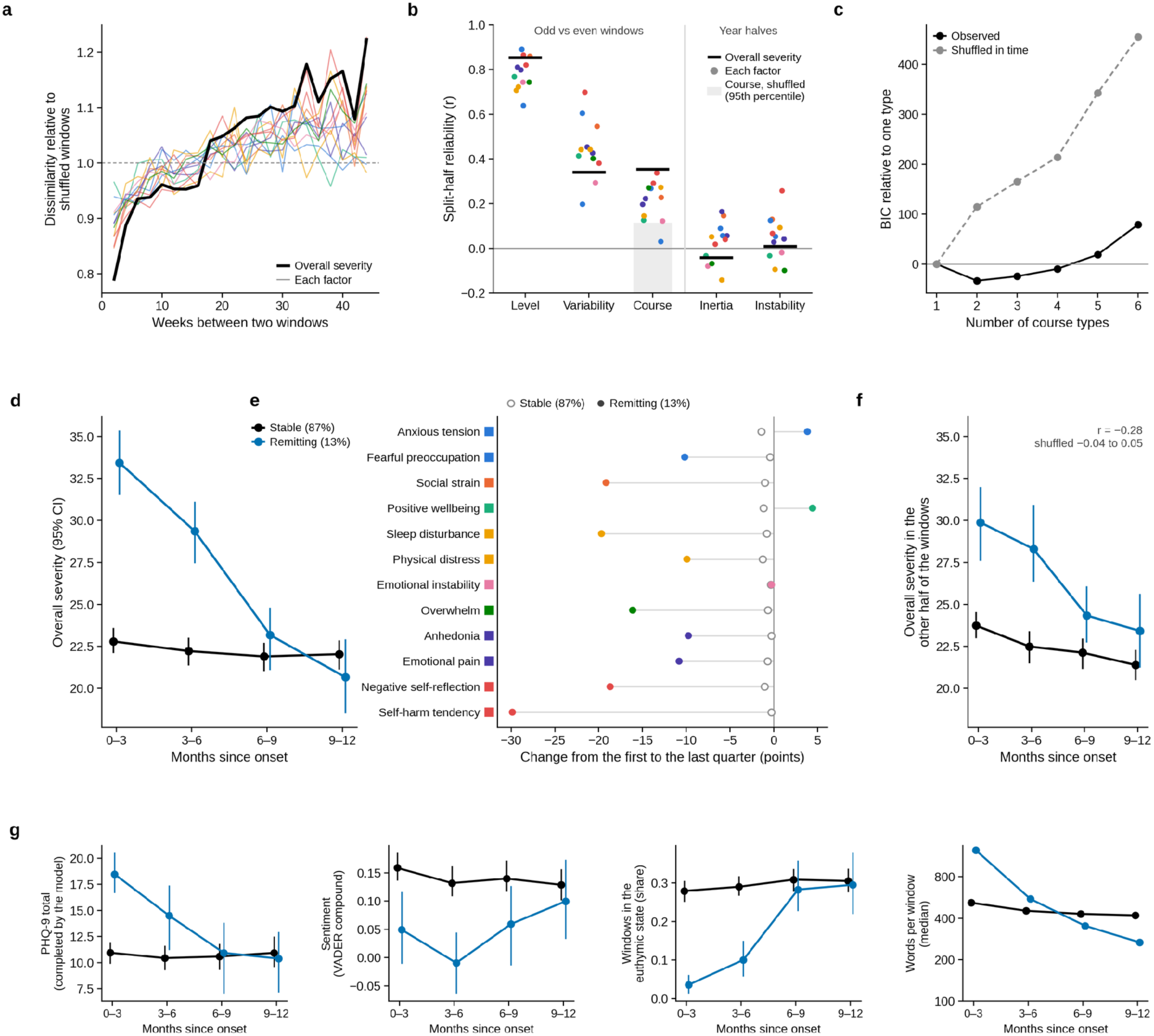
People differ in the course of their symptoms over months. **a**, Dissimilarity of two windows of the same user, half their mean squared difference, by the time between them, relative to the same quantity after shuffling each user’s windows in time (mean of 20 shuffles), for overall severity (the mean of the 11 factors other than Positive wellbeing, black) and for each factor (colors as in Fig. 2). Lags with at least 700 pairs of windows are shown. **b**, Reliability of features of each person for each factor (dots) and for overall severity (bars). Level, variability (standard deviation) and course (slope over the year) were estimated separately from the odd and from the even windows of each user with at least 10 windows (414 users) and correlated across users. The gray band reaches the 95th percentile of the course correlation in 20 shuffles in time. Inertia (lag-1 autocorrelation) and instability (mean squared successive difference) were compared between the first and second halves of the year after regression on level, its square and variability. **c**, BIC of growth mixture models with one to six course types, relative to one type, for the observed windows and for windows shuffled within each user. The log likelihood of users left out of fitting (fivefold cross-validation) was higher than with one type for every number of types on the observed windows (highest with five) and highest with one type on the shuffled windows. **d**, Overall severity by quarter of the year in the two course types, with 95% intervals from 200 bootstrap resamples of users. Percentages give the share of users. **e**, Change of each factor from the first to the last quarter of the year in the remitting (filled) and stable (open) types. **f**, Course in windows not used to assign the type. Models fitted to the other users (five folds) assigned each user with at least 10 windows to a type from the odd windows, and overall severity is shown by quarter in the even windows, and conversely (both directions pooled), with 95% intervals from 200 bootstrap resamples of users. r is the correlation between the log odds of the remitting type from one half and the slope of severity in the other half, with its range in 10 repetitions with windows shuffled in time. **g**, Measures not used to define the types, by quarter of the year. The PHQ-9 total was completed by the same model for the windows that could be linked to their authors, sentiment is the VADER compound score of the posts, the euthymic state is the state PW of Fig. 4 and words per window are medians. Black, stable type. Blue, remitting type.

Interestingly, we found that the longitudinal course was distinct for a remitting group of people compared to a stable majority. A growth mixture model,^52^ which assigns each user to one of several types of course over all 12 factors, selected two types by BIC, against one for shuffled windows (**Fig. 6c**). Most users (87%) followed a stable course, but the other 13%, nearly all from r/depression (95%), entered in a crisis that remitted over six to nine months (**Fig. 6d,e**). Over this remission period, self-harm content, negative self-reflection, social strain, sleep disturbance and overwhelm fell by 16–30 points from the onset, positive wellbeing rose and anxious tension persisted. The PHQ-9 total score completed by the same model fell from 18 to 10 points, against 11 throughout in the stable group (**Fig. 6g**).

The type assigned to a person predicted their course in hold-out windows that were not used to assign it. Models fitted to 80% of users found the remitting type in every fold (correlation with the full model 0.84–0.98). We found that users assigned to the remitting type from their odd or even windows showed a decline in severity (since onset) in the other half (**Fig. 6f**, r = −0.28 between the log odds of the type and the slope in the other half, against −0.04 to 0.05 after shuffling). Finally, these longitudinal courses described a period rather than a trait, as slopes in the first and second halves of the year correlated negatively (r = −0.19).

## Discussion

Here, we introduced TALQ and used it to turn a year of online posts from 491 people into ratings of 52 symptoms every two weeks. The ratings were stable, predicted constructs annotated by humans, agreed with inventories completed by the same model and resolved into 12 replicable mental health factors. Only two lagged relations survived correction for multiple testing, from self-harm content to emotional pain and from sleep disturbance to overwhelm. People differed mainly in which states they occupied, and over months a minority followed a remitting course.

TALQ has properties that are hard to obtain from human raters (**Supplementary Note 2**). The battery can be chosen after data collection, and here posts written years before the 52 symptoms were compiled^6^ were scored against them. The ratings are dense enough to study change within a person, which periodic questionnaires rarely allow,^11,13^ and require no effort from the people assessed. Because the model has open weights, TALQ can run locally, keeping sensitive text within the institution that holds it. The ratings remain inferences about expressed mental states, not diagnoses, and they inherit the priors of the model, the conventions of each community and what people choose to write.

The 12 factors recovered the main symptom domains of depression and anxiety without training on clinical labels. The separation of anxious tension from fearful preoccupation parallels the distinction between distress and fear in the Hierarchical Taxonomy of Psychopathology (HiTOP), which separates generalized anxiety from panic.^53^ Social strain has no counterpart among the diagnostic criteria but is central to interpersonal theories of suicide,^54^ and it accompanied self-harm content in the most distressed states and regimes.

The lagged relation from sleep disturbance to overwhelm agrees with clinical observations. Prior studies show that insomnia predicts the onset of depression,^55^ poor sleep is followed by worse mood the next day,^56^ and treating insomnia reduces depressive and other psychiatric symptoms,^57^ which makes sleep a modifiable entry point. Our data add overwhelm, a domain of strained effort, poor concentration and agitation, to this sequence.

Our finding that self-harm-related content predicted greater emotional pain two weeks later may seem counterintuitive, given theories that emphasize psychological pain as a driver of suicidal ideation.^46^ However, the relationship may be bidirectional. Intensive longitudinal studies have found that suicidal ideation can also predict subsequent hopelessness, negative affect, and psychological pain over hours to days.^47–49^ Our findings are consistent with this possibility at a longer interval, although they reflect language-derived symptom factors rather than clinically assessed suicidal ideation or psychological pain. Together, these observations suggest that the temporal relationship between suicidal thinking and emotional distress may be more complex than a unidirectional model would suggest. Future studies examining both directions across multiple timescales could help clarify these relationships.

The weak dynamics between consecutive windows may partly reflect our design. Windows of two weeks average over the faster fluctuations that experience sampling captures,^11,58^ and autoregressive estimates from short panels are biased toward zero,^59^ a bias that the jackknife reduces but may not remove.^45^ These results are therefore not evidence against symptom networks at shorter time scales. Slower change was more informative, whereas inertia and instability were not reliable, in line with evidence that such indices add little to the mean and variability of affect.^60^ Monitoring between visits may therefore be most informative about the states a person tends to occupy and the direction in which their symptoms move over months.

The longitudinal analysis of symptom factors points to a clinically distinct group. About 13% of the users in our cohort joined during a crisis dominated by self-harm content, self-criticism and social strain that resolved over six to nine months while anxious tension persisted. Such course types are familiar from cohort studies of depressive symptoms^61^ and could guide care, for example by attending to residual anxiety once a crisis resolves. Because mixtures of course types can approximate a continuum^62^ and assignments agreed weakly between halves, membership is better reported as a probability than as a label.

This study has several limitations. First, the cohort of highly active members of two communities writing in English between 2009 and 2018 does not represent people with depression or anxiety in general. Second, the ratings came from one model, prompt and battery, and a newer model rated only the validation posts and a few cohort windows. Language models can also carry social biases that differ across groups of writers,^63^ which we could not assess without demographic information. Third, the confidence filter removed windows with less text more often, and windows without posts were treated as missing, although not posting may be informative. Fourth, each year began when posting became sustained, so courses describe the year after people began to seek support. Finally, factors, states and course types are simplifications that depend on modeling choices, and Granger predictive relations should not be interpreted as causal effects. Our future work also includes assessing the robustness of the framework against noisy textual data^64^ such as electronics health records, as well as the safety of the model against adversarial attacks.^65,66^

TALQ can turn naturalistic language into dense, reproducible and replicable ratings of mental states. At a resolution of two weeks, little evidence remains for coupled symptom dynamics beyond two lagged relations, whereas over months people differed in the course of their symptoms. Validation against clinician ratings should precede clinical uses, such as summaries of the interval between visits from text that patients choose to share, which also require attention to consent, privacy and the risk of overinterpretation.

## Methods

### Data source

We analyzed the ConvoKit corpora of r/depression and r/Anxiety,^67^ which contain every post and comment (together, utterances) from the creation of each community to 31 October 2018. They comprise 4,547,682 utterances from 549,783 account names, including “[deleted]”. The archives were used as distributed, and their SHA-256 checksums are listed in the code repository. The study used only publicly posted, previously collected data. We did not interact with any user and made no attempt to identify users.

### Cohort and observation windows

For every account with at least 20 utterances (33,520 accounts), we estimated a daily posting rate by summing Epanechnikov kernels centered on each utterance, each extending 28 days to either side. A day counted as active if the rate was at least 0.2 utterances per day. We retained the accounts with at least 360 active days, which need not be contiguous, and excluded one of them, AutoModerator, a moderation bot. This left 491 users. Onset was the first active day. Each user’s first year after onset was divided into 27 consecutive windows of 14 days, the first beginning about one day before onset. Each window contained every post and comment the user wrote in either community. Window boundaries were defined in Pacific time (America/Los_Angeles). Windows without utterances were not scored, which left 9,886 windows (median 22 per user) with 132,765 utterances, of which 6.2% were posts. Windows contained a median of 7 utterances (interquartile range (IQR) 3–15) and 417 words (IQR 147–1,023).

### Language model questionnaire

In TALQ, each window was rated on each of the 52 symptoms compiled by Fried^6^ (**Supplementary Table 1**). The system message cast the model as a mental health professional assessing a client from the Reddit posts of the last two weeks. It listed the utterances of the window and told the model to rely only on evidence in the posts and to answer in JSON. The user message gave the title of the symptom, a short description for 40 of the 52 symptoms and a rubric from 0 (“symptom is entirely absent”) to 100 (“symptom is present every day”). The model returned a reasoning of at most four sentences, an integer rating and an integer confidence from 0 to 100 that the rating was supported by evidence (prompts in **Supplementary Note 1**).

Ratings were generated with Llama-3.1-8B-Instruct^33^ quantized to 4 bits in the NormalFloat format (NF4) with bitsandbytes and transformers 4.44.2. Sampling used the default settings of the checkpoint (temperature 0.6, nucleus sampling with p = 0.9 and sampling from the 50 most likely tokens) with at most 384 new tokens. Each window and symptom was rated five times (2,570,360 requests). Responses were stored for 2,545,783 requests, and the remaining 0.96% were treated as missing. Responses were parsed as JSON, and 93.0% were valid JSON. For the others, we extracted an embedded JSON object or, failing that, read the rating and confidence with regular expressions (6.9% of responses). Responses that could not be parsed and values outside the scale were treated as missing (3,779 responses, 0.15%, **Extended Data Fig. 2**).

### Symptom profiles

Ratings and confidences were averaged over the available replicates of each window and symptom. We kept users with at least 200 rated window–symptom pairs, and windows in which all 52 symptoms were rated and the mean confidence exceeded 40 for every symptom (7,094 windows from 481 users). Replicate reliability was ICC(1), the intraclass correlation of single ratings across replicates.^68^

### Latent mental factors

The matrix of windows by symptoms was decomposed by nonnegative matrix factorization^69^ in scikit-learn,^70^ with coordinate descent, Frobenius loss and a tolerance of 10⁻⁵. We assessed dimensionality with staNMF,^40^ the mean pairwise Amari distance between solutions from different random initializations, in which K randomly chosen symptom columns served as initial window scores. A broad sweep used 20 initializations for each K from 3 to 25, and a fine sweep used 100 for each K from 11 to 14, around the dimensionality at which factors were most interpretable in preliminary fits, and for K from 17 to 19, around the lowest instability of the broad sweep above 10 factors. We used K = 12, the most stable dimensionality between 11 and 14. The final model was the fit with the lowest error among 100, and scores were rescaled to range from 0 to 100 by the maximum of each factor. To describe the alternatives, the fits with the lowest error at K = 10 (10 initializations) and K = 18 (100 initializations) were matched to the main factors by Tucker congruence (**Extended Data Fig. 5e**). Llama-3.1-8B proposed candidate labels from the most heavily weighted symptoms of each factor. The authors chose the labels and mapped the factors to DSM-5-TR criteria^42,43^ (**Table 1**).

Replicability was assessed with Tucker’s congruence^44^ after optimal matching of factors. We refitted the model in the two halves of 20 random splits of users, compared with each other, at confidence thresholds of 30 and 50, compared with the main loadings, and in the users who wrote most of their utterances in r/depression and in those who wrote most in r/Anxiety, also compared with each other. Refits at other thresholds and in each community used the lowest error of five initializations, and the congruence between the communities was compared with that between ten random splits of all users into two groups of the same sizes.

### Psychometric properties

Internal consistency was the weighted Cronbach’s α^71^ of the composite y = Σᵢ wᵢxᵢ weighted by the factor loadings, α = [k_eff/(k_eff − 1)][1 − Σᵢ var(wᵢxᵢ)/var(y)] with k_eff = (Σᵢ wᵢ)²/Σᵢ wᵢ², which reduces to k/(k − 1) for equal weights. Because the weight of each factor is concentrated on a few symptoms (k_eff 6.3–21.3), we also report ordinary α over the items that carry 50% of the weight (**Extended Data Fig. 6**). Item discrimination was the correlation of each item with the rest of the composite, averaged over the items that carry 50% of the weight of each factor. Both properties were also estimated within strata of model confidence. Intervals for α came from 200 bootstrap resamples of users.

### Factor dynamics within individuals

Lagged dependencies were estimated with a panel vector autoregression^72^ on pairs of windows of the same user exactly 14 days apart, with scores centered within user.^73^ The lag order was chosen by BIC among orders 1 to 3 on a common sample. Centering short panels within users biases autoregressive coefficients toward zero,^59^ by about 0.1 in simulations matched to our panel. We therefore used the half-panel jackknife,^45^ A = 2Â − (Â₁ + Â₂)/2, where Â₁ and Â₂ are estimated on the first and second halves of each user’s span. A bootstrap over users (500 resamples) gave Wald tests for each ordered pair, with FDR correction over the 132 pairs,^74^ and confidence bands for impulse responses to a shock of 50 points and step responses to a sustained change of 25 points. We also simulated 20 panels that kept the observation pattern of each user, the autoregressive coefficient of each factor and the covariance of the innovations, but had no lagged effects between factors. In these panels the test rejected 4.3% of pairs at P < 0.05, with 0.15 FDR discoveries on average.

### Discrete mental states

Factor vectors were embedded with uniform manifold approximation and projection (UMAP)^75^ for visualization (20 neighbors, minimum distance 0.08 and correlation metric), and trustworthiness (k = 20) measured the faithfulness of the embedding. They were clustered with a truncated Dirichlet process Gaussian mixture fitted by variational inference^76^ in scikit-learn (100 components, full covariances and 10 initializations) at 14 hyperparameter settings. Across settings, the prior on weight concentration ranged from 10⁻⁸ to 10⁻¹⁴, the prior on mean precision from 0.001 to 3 and the regularization of covariances from 10 to 150. The most populated components that together covered 90% of windows were retained. The setting was chosen by BIC, computed with the parameter count of a Gaussian mixture with full covariances and the retained components, and with the number of windows as the sample size. The sweep ran from the least regularized setting (step 0) to the most regularized (step 13). BIC was lowest at step 5, which gave 11 states, whereas AIC and AICc favored the most complex setting (step 0, 39 states) (**Extended Data Fig. 8**).

States were labeled by the factors whose centroid exceeded the mean over states by at least one standard deviation, with at most two factors per label and the highest factor given in parentheses when no factor qualified. For the sensitivity analysis, we used the adjacent, more regularized step (step 6, eight states).

### Dynamical regimes

State sequences of users with at least 10 confident windows and no gap longer than 57 days (249 users and 4,436 windows) were modeled as a mixture of first-order Markov chains.^50^ The mixture was fitted by expectation–maximization with hard assignments, with occupancy as the distribution of initial states, transitions across gaps counted as single steps and 1,000 random restarts (10,000 for the final model). The number of regimes was chosen by AICc.^51^

We evaluated the regimes in five ways. First, we computed the log likelihood per window of users left out of fitting under fivefold cross-validation over users, both for Markov mixtures (expectation–maximization with soft assignments, Dirichlet smoothing and transitions that respect gaps) and for mixtures of multinomial distributions that model occupancy only. Second, a null model permuted each user’s windows in time 200 times, which preserves occupancy, and was applied to the Markov advantage and to state persistence. Third, we measured the agreement of the regime labels with labels from soft assignments, from occupancy mixtures and from 50 bootstrap refits with the adjusted Rand index.^77^ Fourth, to assess stability within people, models fitted on 80% of the users classified the first and second halves of the year of each remaining user, and the agreement between halves was compared with a permutation null. Fifth, we repeated these analyses with eight states.

Symptom burden was the share of each user’s windows at or above the 90th percentile of Regime 1. A Gaussian hidden Markov model^78^ with the same number of states was also fitted to the factor scores of contiguous runs of windows, with full covariances and three initializations (**Extended Data Fig. 10**).

### Course of symptoms over months

Overall severity was the mean of the 11 factors other than Positive wellbeing. Slow change was described by the semivariogram, half the mean squared difference between two windows of the same user as a function of the number of windows between them, divided by its mean over 20 shuffles of each user’s windows in time. For the 414 users with at least 10 confident windows, the level (mean), variability (standard deviation) and course (least-squares slope over the year) of each factor and of overall severity were estimated separately from the odd and from the even windows of each user, two interleaved halves that cover the same year, and correlated across users, with a null for course from 20 shuffles in time. Inertia, the lag-1 autocorrelation over exactly adjacent windows, and instability, the log mean squared successive difference, were compared between the first 13 and the last 14 windows of the year after regression on level, its square and variability. Course types were estimated with a growth mixture model.^52^ Each user belonged to one of K types, each with its own linear and quadratic trend in time for every factor, around a random intercept per factor with a full covariance, and windows shared a full residual covariance across the 12 factors. The model was fitted by expectation–maximization with 10 random starts for K = 1 to 6, on the observed windows and on windows shuffled within each user. K was chosen by BIC with the number of users as the sample size, and the log likelihood of users left out of fitting was computed under fivefold cross-validation over users. Models fitted to 80% of users classified the odd and the even windows of each remaining user with at least 10 windows. Agreement between the two classifications was summarized by Cohen’s κ and by the correlation of the log odds of the smaller type, and the prediction of course by the correlation of the log odds from one half with the slope of overall severity in the other half, each against 10 repetitions with windows shuffled in time. The model was also refitted in the users who wrote mostly in r/depression. Types were described by quarter of the year (windows 0–6, 7–13, 14–20 and 21–26), with intervals from 200 bootstrap resamples of users, by the factors, the inventories completed by the same model, the VADER compound score^79^ of the posts and the share of windows in the euthymic state.

### Validation against human annotations

The TALQ prompt was applied to single posts, each presented as a window containing one post, from three Reddit datasets with labels assigned by human annotators. The first dataset labeled two interpersonal risk factors, thwarted belongingness and perceived burdensomeness,^35,54^ the second low self-esteem^36^ and the third depression severity on four levels (minimum, mild, moderate and severe).^37^ Before any model output on these posts existed, each label was mapped to battery symptoms, and the SHA-256 hash of the mapping was recorded (**Supplementary Table 2**). For example, thwarted belongingness was mapped to loneliness, “other people dislike me”, “other people were unfriendly to me” and interpersonal sensitivity, and severity to the 20 items that cover criterion A of DSM-5-TR. Three control symptoms without a conceptual link to the labels (initial insomnia, decreased appetite and gastrointestinal problems) were also specified in advance. The mapping determined which symptoms were scored but was not used by the predictive models.

To test the scoring system in a deployment that keeps text on local hardware, we ran the validation on a single computer with an Apple M5 processor, with Llama-3.1-8B-Instruct quantized to 4 bits (Q4_K_M) and served locally by llama.cpp. Prompts were rendered and sampled as in the cohort annotation (**Supplementary Note 1**), with five sample per request. A reduced protocol was fixed in advance. It comprised balanced random samples of 300 posts per dataset, scored on the mapped and control symptoms (12 symptoms for the interpersonal labels, 8 for low self-esteem and 20 for severity, 60,000 generations in total).

For each label, the ratings of all symptoms scored on its dataset were combined by logistic regression with an L1 penalty (lasso),^38^ or for severity by linear regression of the level with an L1 penalty, with the penalty chosen by fivefold cross-validation within the training posts, and by a neural network with one hidden layer of 16 units and an L2 penalty chosen in the same way. Ratings were standardized and missing ratings replaced by the median, both estimated on the training posts. Predictions came from fivefold cross-validation over posts, stratified by label and repeated ten times, and were averaged over repetitions. Prediction was summarized by the AUROC, for severity of severe against minimum depression, and for severity also by Spearman ρ with the level. Intervals and paired differences came from 2,000 bootstrap resamples of posts. The gain from averaging was the paired difference in AUROC between the lasso on the mean of five ratings and the lasso on the first rating alone. A symptom was taken to carry the prediction when the lasso kept it in at least 80% of the 50 training folds.

The comparators were the best single symptom, chosen within the same training folds, nine lexicon and length features and a supervised reference. The features were the negativity and compound scores of VADER,^79^ the Empath categories negative emotion, sadness, shame and suffering,^80^ the rates of first person singular pronouns^81^ and of absolutist words,^82^ and post length, combined by the same lasso as the ratings. The supervised reference was logistic regression with L2 regularization on term frequency–inverse document frequency (TF-IDF) features, with predictions from fivefold cross-validation over each full dataset. To measure the effect of quantization and runtime, the calibration windows, drawn at random from cohort windows short enough for a context of 4,096 tokens, were rated on eight symptoms and compared with the cohort ratings (**Extended Data Fig. 3c,d**). The ceiling for a single sample is the agreement of one cohort sample with the mean of the other four.

### Agreement with standard inventories completed by the same model

The same model also answered the BDI-II^16^ and the PHQ-9^15^ for the same windows, in the first person. For each of the 30 items it chose an option scored from 0 to 3, five times per item (1,479,190 responses, each scored by the digit of the chosen option). The inventory outputs were keyed by the start of the window only. Because window starts lie on a grid of 14 days shared across users, we linked a window to its author only when a single cohort member had a window starting at that time. This linked 1,720 windows from 238 users, of which 1,681 windows from 235 users had complete battery ratings. Inventory items were mapped to battery items in advance (**Supplementary Table 3**). We report the Spearman correlation of each total with the mean of the mapped battery items, pooled and within person, with intervals from a bootstrap over users. We also report the correlation of each item with its mapped items (convergent) and its median correlation with the other items (discriminant).

### Additional language models

Qwen3.6-35B-A3B,^39^ released in 2026, has 35 billion parameters, of which 3 billion are active for each token. It rated the validation posts and calibration windows of the reduced protocol once (300 posts per dataset) on the same computer, served by llama.cpp with weights quantized to about 4 bits (UD-IQ4_XS). It used its own chat template with its thinking mode switched off and the same sampling settings. Because it rated each post once, it was compared with the first of the five Llama-3.1-8B-Instruct ratings, and differences in AUROC between the two models were estimated from 2,000 paired bootstrap resamples of posts. Dependence on text length was summarized by the Spearman correlation between the mean rating of each post and its log word count.

## Statistics and software

Unless stated otherwise, intervals are 95% percentile bootstrap intervals over users, tests are two-sided and multiple comparisons are controlled by the Benjamini–Hochberg procedure. Analyses used Python 3.11 with numpy, pandas, scipy, scikit-learn,^70^ statsmodels,^83^ umap-learn^75^ and hmmlearn. All results and figures can be regenerated from the public corpora and the stored model outputs by numbered scripts with fixed seeds.

## Data availability

The ConvoKit corpora are publicly available at https://convokit.cornell.edu. The validation datasets annotated by humans are available at (1) https://github.com/am-shb/irf-acl, (2) https://github.com/drmuskangarg/LoST, and (3) https://github.com/usmaann/Depression_Severity_Dataset.^35–37^

## Code availability

All code for data acquisition, cohort selection, prompt construction, annotation, analyses and figures is available at https://github.com/abbasilab/TALQ.

## Data Availability

https://convokit.cornell.edu

https://github.com/am-shb/irf-acl

https://github.com/drmuskangarg/LoST

https://github.com/usmaann/Depression_Severity_Dataset

https://github.com/abbasilab/TALQ

## Acknowledgements

The authors thank A. Taylor, G. Cui, N. Donovan, R. P. Xian, M. Bijanzadeh, C. Taylor, D. Willoughby, D. H. Hadjiabadi, A. Gallagher, V. Sohal and C. Kirst for feedback, and D. G. K. Walker for contributions to the genesis of the project. MEL would like to acknowledge support from NIH R01NS099254 and the UCSF Weill Fellowship. RAA would like to acknowledge support from the UC Noyce Initiative. During this work, the authors used LLMs as an aid in writing analysis code, searching the literature and editing the manuscript text. The authors directed this use, reviewed and verified all code, results, references and text, and take full responsibility for the content of the publication.

## Author contributions

RAA and MEL conceived the study. MEL curated the data. RAA and MEL conceptualized the analytical framework and performed the analysis. RAA, MEL, and JW interpreted the findings. KEP contributed to funding and resources used to support the study. RAA supervised the study. RAA and MEL wrote the manuscript. All authors reviewed and revised the manuscript.

## Extended Data figure legends

**Extended Data Fig. 1.**
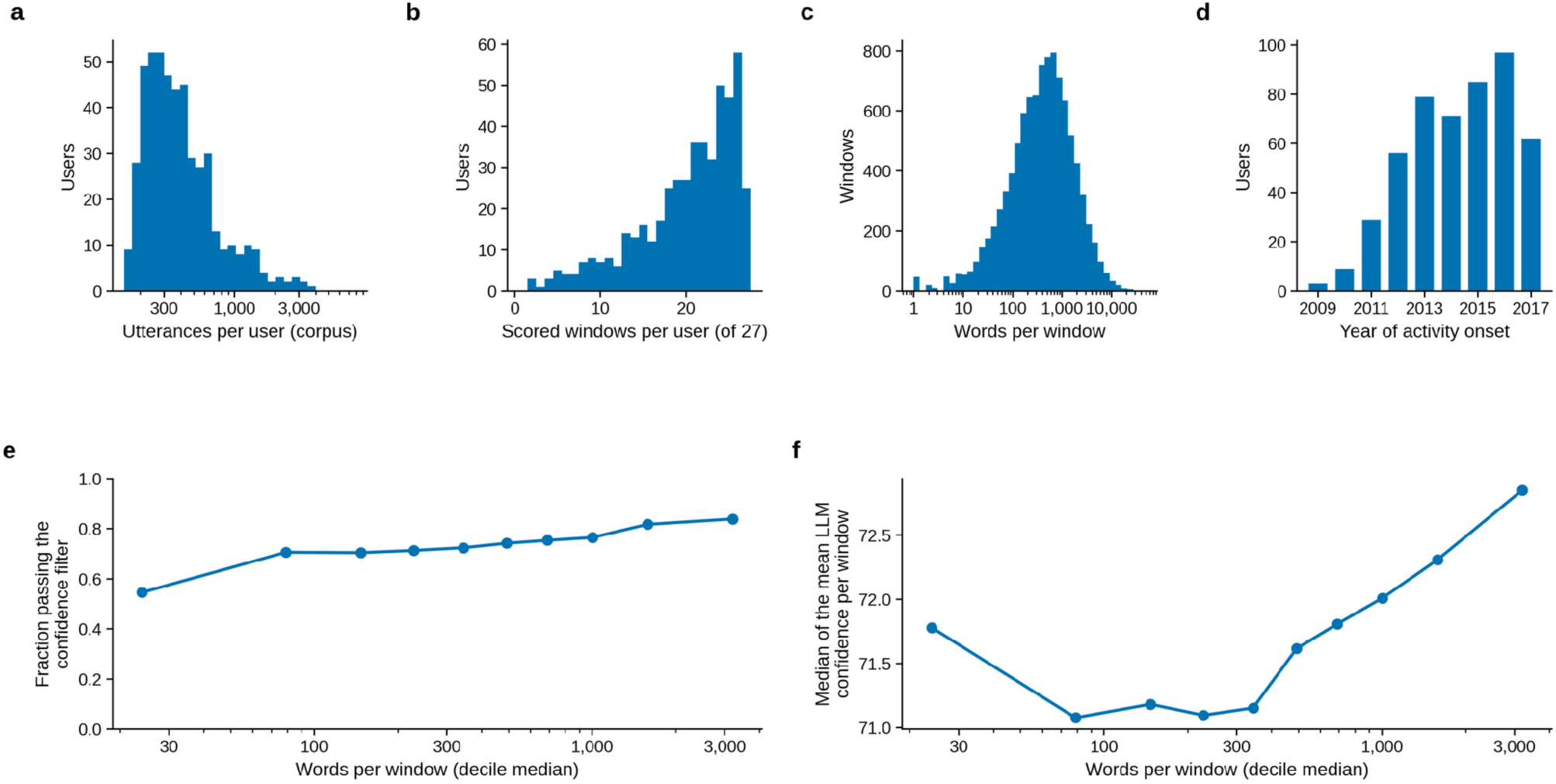
Cohort, text volume and the confidence filter. **a**, Utterances per user in the two corpora. **b**, Scored windows per user, out of 27. **c**, Words per scored window. **d**, Year of onset of sustained activity. **e**, Fraction of complete windows that passed the confidence filter (confidence above 40 for all 52 symptoms), by decile of words per window. **f**, Median of the mean model confidence of each window, by decile of words per window (Spearman ρ between confidence and word count 0.11 across windows).

**Extended Data Fig. 2.**
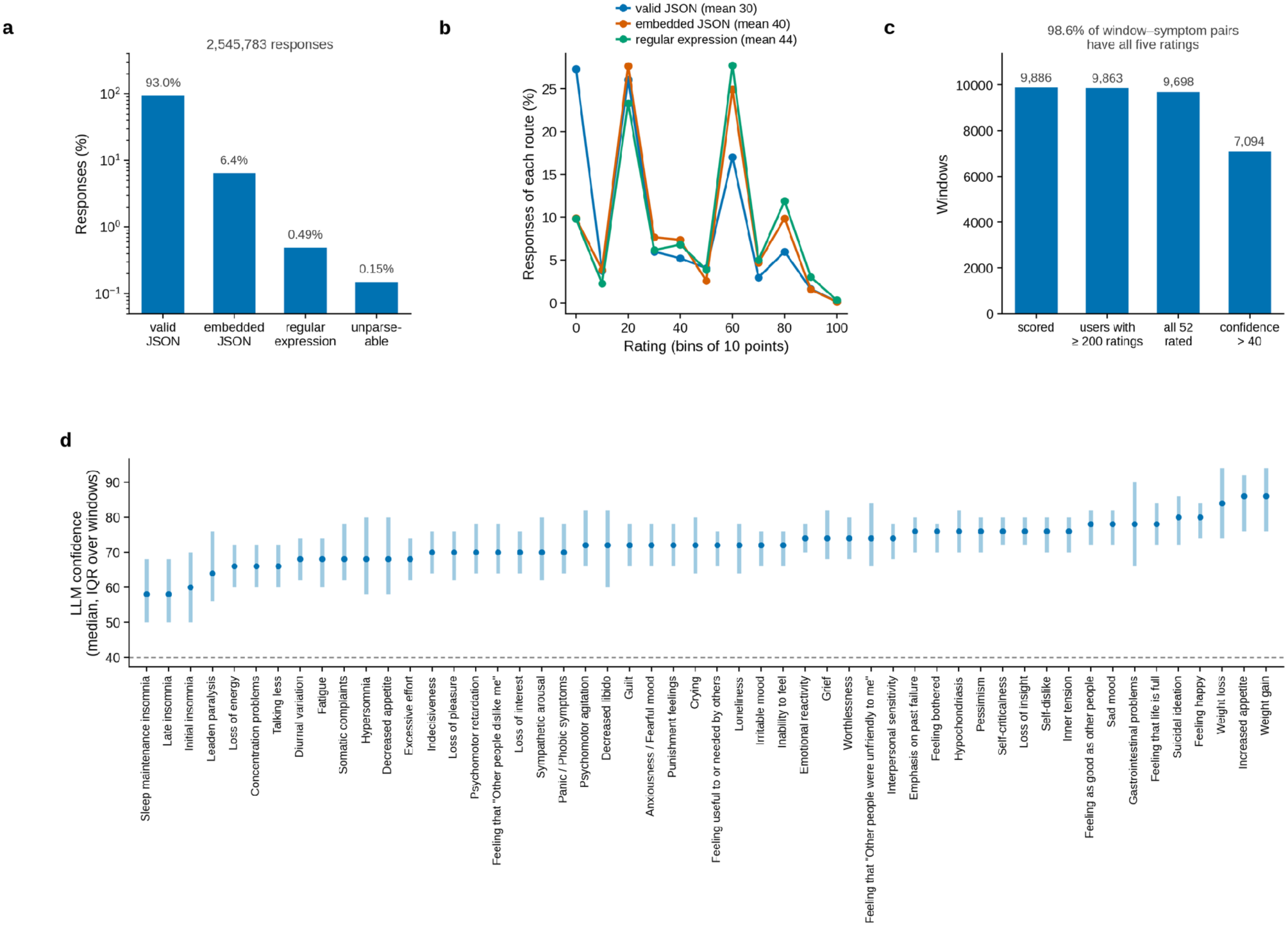
Response parsing and window flow. **a**, Parse route of all responses for the 491 users. Responses were valid JSON, contained a JSON object embedded in other text, were read by regular expressions or could not be parsed and were treated as missing. **b**, Distribution of ratings by parse route, in bins of 10 points. Ratings recovered from embedded JSON or by regular expressions were higher on average because these routes were more common for window–symptom pairs with high ratings (2.6% of responses in the lowest and 10.4% in the highest quintile of the mean rating of the pair). Within the same window and symptom, ratings differed little by route (mean difference 0.6 points). **c**, Windows remaining at each step, from all scored windows to windows of users with at least 200 rated window–symptom pairs, to windows with all 52 symptoms rated and to windows with confidence above 40 for every symptom. The share of all requested window–symptom pairs with five valid ratings is given above the panel. **d**, Median model confidence per symptom, with the interquartile range over windows. The dashed line marks the filter threshold of 40.

**Extended Data Fig. 3.**
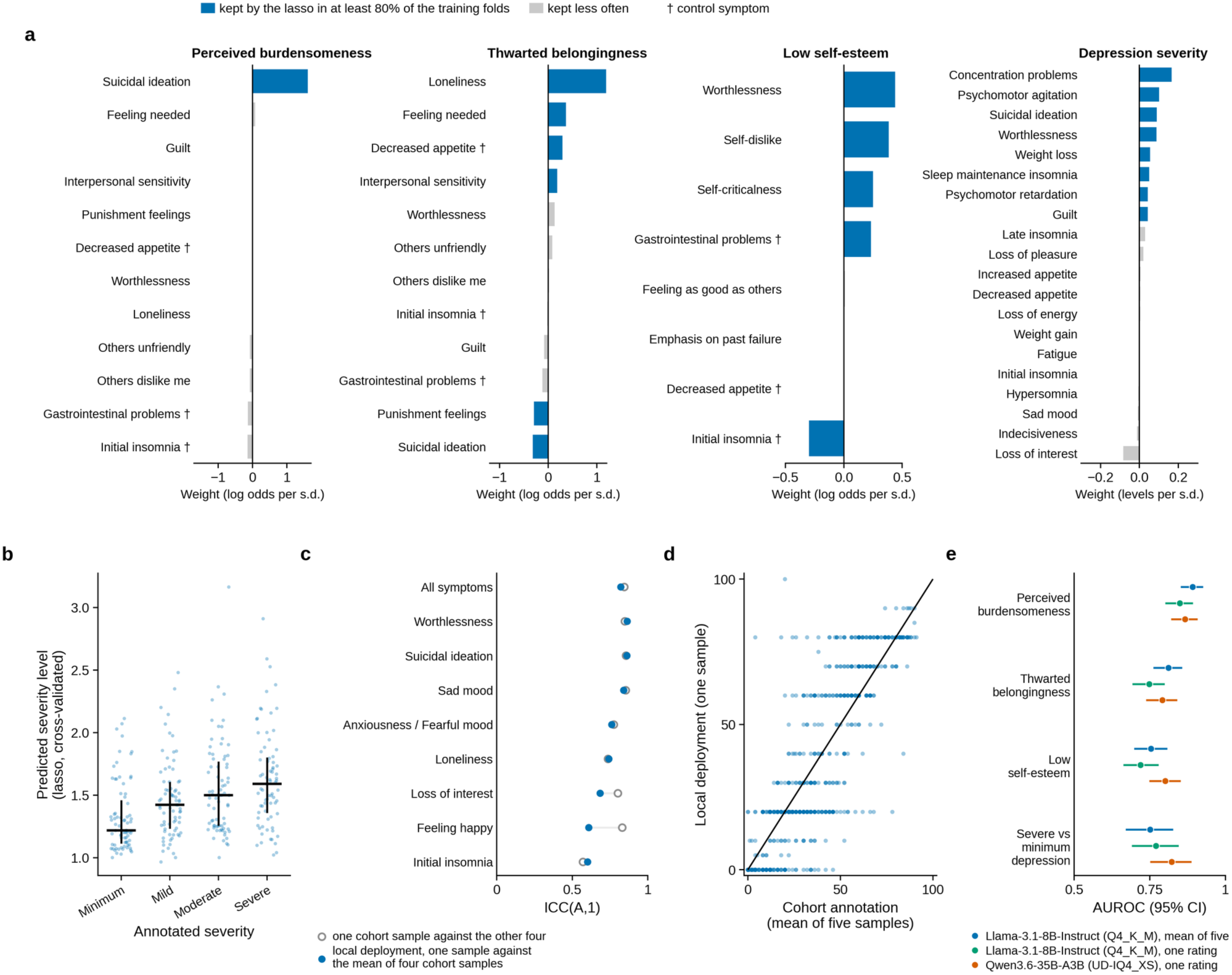
Prediction of human annotations in detail, from the reduced protocol run with a local deployment of the rater. Panels **a** to **d** show Llama-3.1-8B-Instruct, the annotating model of the main analyses. **a**, Weight of each scored symptom in the lasso that predicts each label from the mean of five ratings per post and symptom, the mean standardized coefficient over the 50 training folds, in log odds per standard deviation of the rating for the binary labels and in levels per standard deviation for severity. Blue bars mark symptoms that the lasso kept in at least 80% of the training folds, and daggers mark the three control symptoms. Of these, the lasso kept decreased appetite for thwarted belongingness, and gastrointestinal problems and initial insomnia, the latter with a negative weight, for low self-esteem. Long names are abbreviated as in Fig. 1b. **b**, Cross-validated prediction of the severity level by annotated level (median and interquartile range, Spearman ρ between prediction and level 0.33, 95% CI 0.23–0.43). **c**, Agreement of one sample of the local deployment with the mean of four samples of the cohort annotation for the same windows, per symptom (ICC(A,1), filled). Open markers show the agreement of one sample of the cohort annotation with the mean of the other four, the ceiling for a single sample. **d**, Rating of the local deployment against the mean of the five samples of the cohort annotation, for all eight calibration symptoms. **e**, Prediction of each label by the lasso for Llama-3.1-8B-Instruct with the mean of its five ratings (blue) and with its first rating alone (green), and for Qwen3.6-35B-A3B, which rated each post once (vermillion), as the AUROC with 95% bootstrap intervals, comparing severe with minimum depression for severity. Across the 12,640 requests that both models answered once, Qwen3.6-35B-A3B declined none, against 1.1% for Llama-3.1-8B (4.6% of requests about posts labeled for perceived burdensomeness), and returned valid JSON for 98% of requests, against 82%. Ratings of both models increased with post length (Spearman ρ between the mean rating of a post and its log word count 0.33 for Llama-3.1-8B and 0.40 for Qwen3.6-35B-A3B).

**Extended Data Fig. 4.**
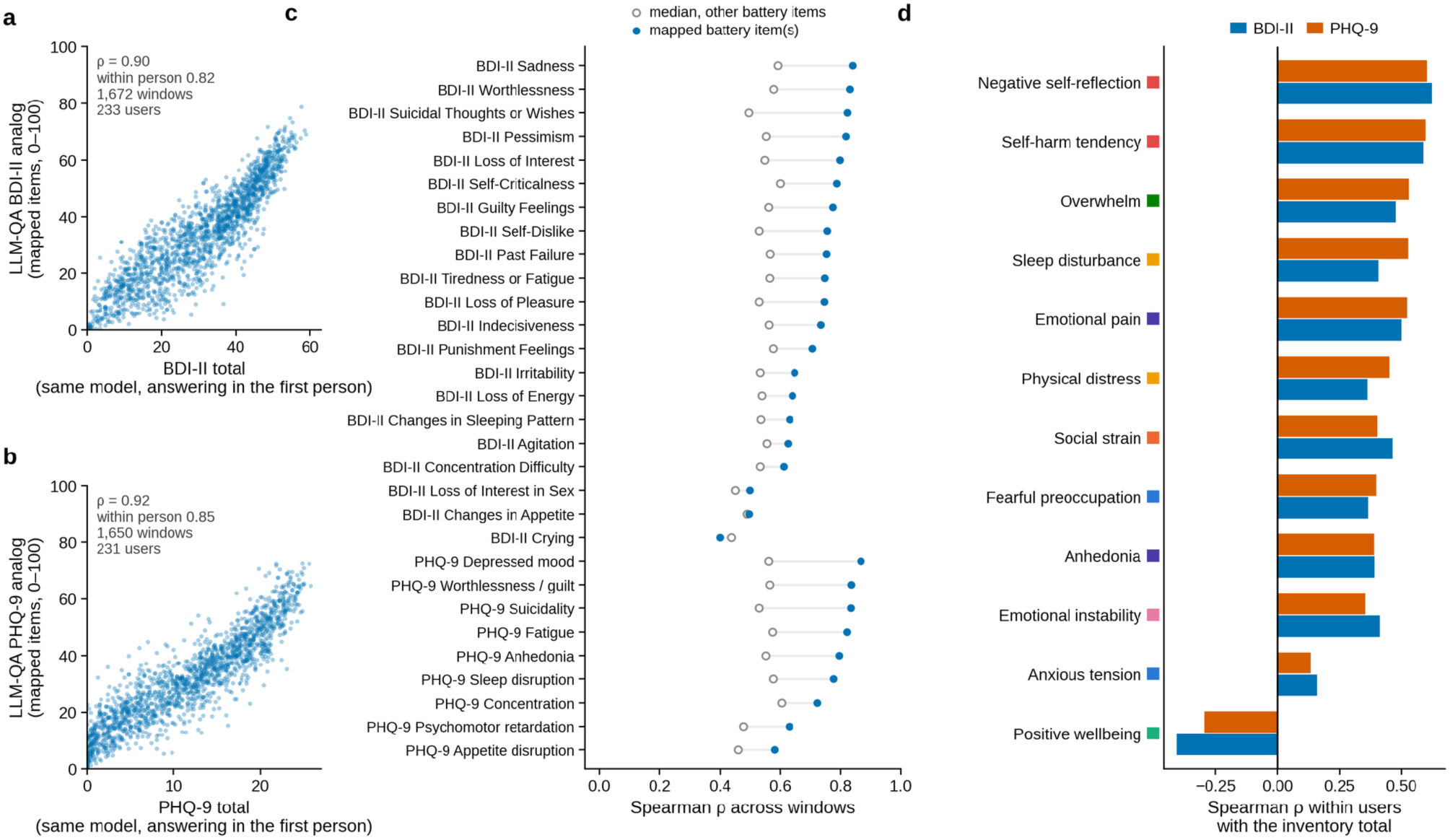
Agreement of the TALQ battery with two standard inventories completed by the same model. **a**, BDI-II total (21 items, each the mean of five samples, answered in the first person) against the TALQ analog of the BDI-II, the mean of the mapped battery items (**Supplementary Table 3**). The panel includes the windows whose author could be identified and whose battery and inventory ratings were complete (numbers in the panel). **b**, The same for the PHQ-9. **c**, Spearman correlation of each inventory item with its mapped battery item or items (filled) and its median correlation with the other battery items (open). **d**, Spearman correlation within users of each factor score with the BDI-II and PHQ-9 totals, after centering within each user, for users with at least three linked confident windows. Colored squares mark the clinical family of each factor as in Fig. 2.

**Extended Data Fig. 5.**
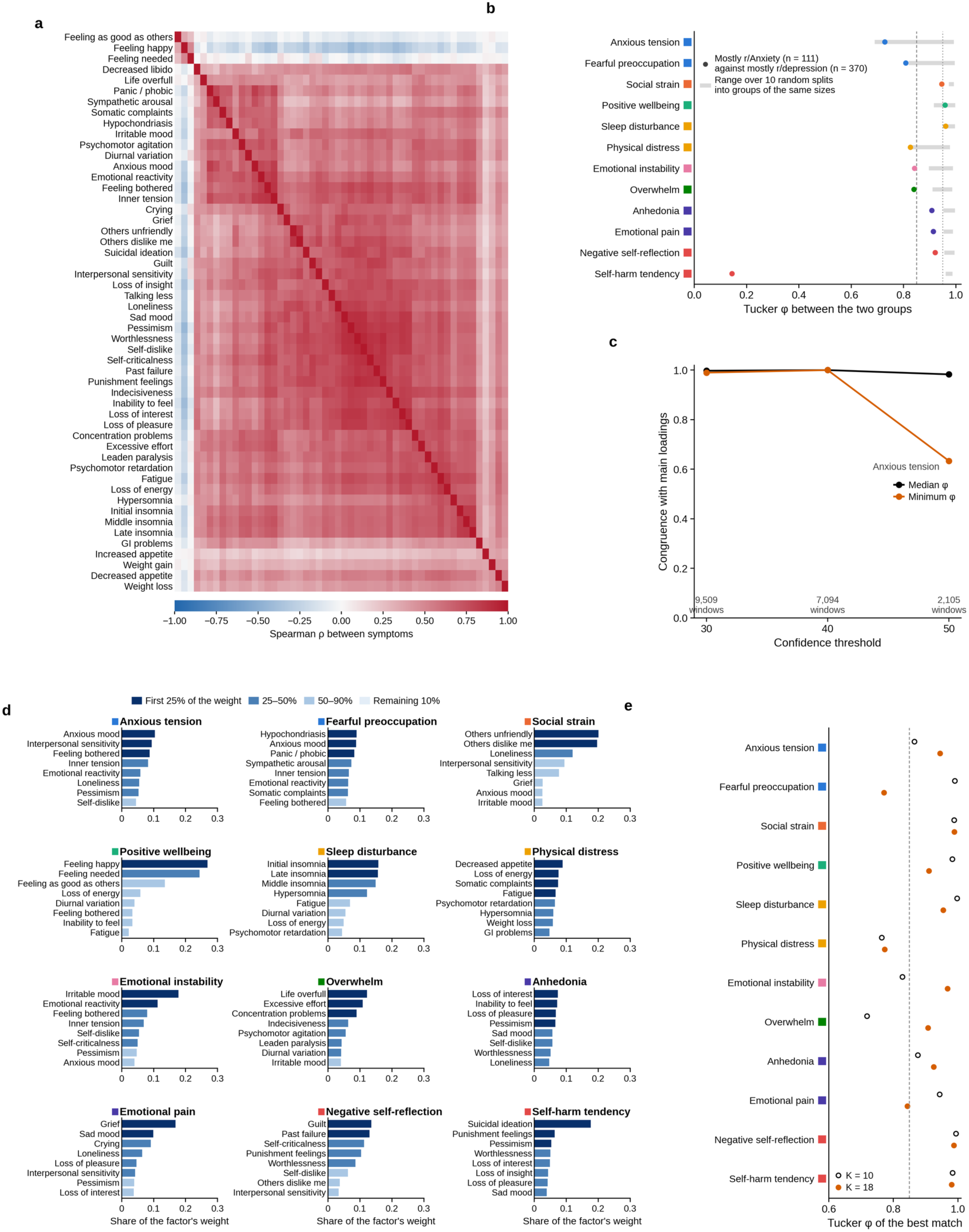
Symptom correlations, replicability of the factor structure and alternative numbers of factors. **a**, Spearman correlations between the 52 symptom ratings across confident windows, ordered by hierarchical clustering. **b**, Tucker congruence between the factors refitted separately in the users who wrote most of their utterances in r/depression and in those who wrote most in r/Anxiety (dots). Bars show the range over ten random splits of all users into two groups of the same sizes. Dashed and dotted lines mark 0.85 and 0.95. **c**, Congruence with the main loadings of factors refitted at confidence thresholds of 30, 40 and 50 (lowest error of five initializations), shown as the median and minimum over factors. The least similar factor is named when its congruence is below 0.85, and the number of windows retained is shown below each threshold. **d**, Composition of each factor. Bars show the share of the factor’s weight carried by its eight most heavily weighted symptoms, shaded by the cumulative share of the weight they complete (first 25%, 25–50%, 50–90% and the remaining 10%). **e**, Tucker congruence between each main factor and its best match among the factors of the best fit with 10 factors (open) and with 18 factors (filled). With 10 factors, Physical distress merged with Anhedonia and Overwhelm with Emotional instability. With 18 factors, five factors placed more than 30% of their weight on a single symptom (hypochondriasis, feeling needed, feeling happy, diurnal variation and grief), against none of the main factors. Colored squares mark the clinical family of each factor as in Fig. 2.

**Extended Data Fig. 6.**
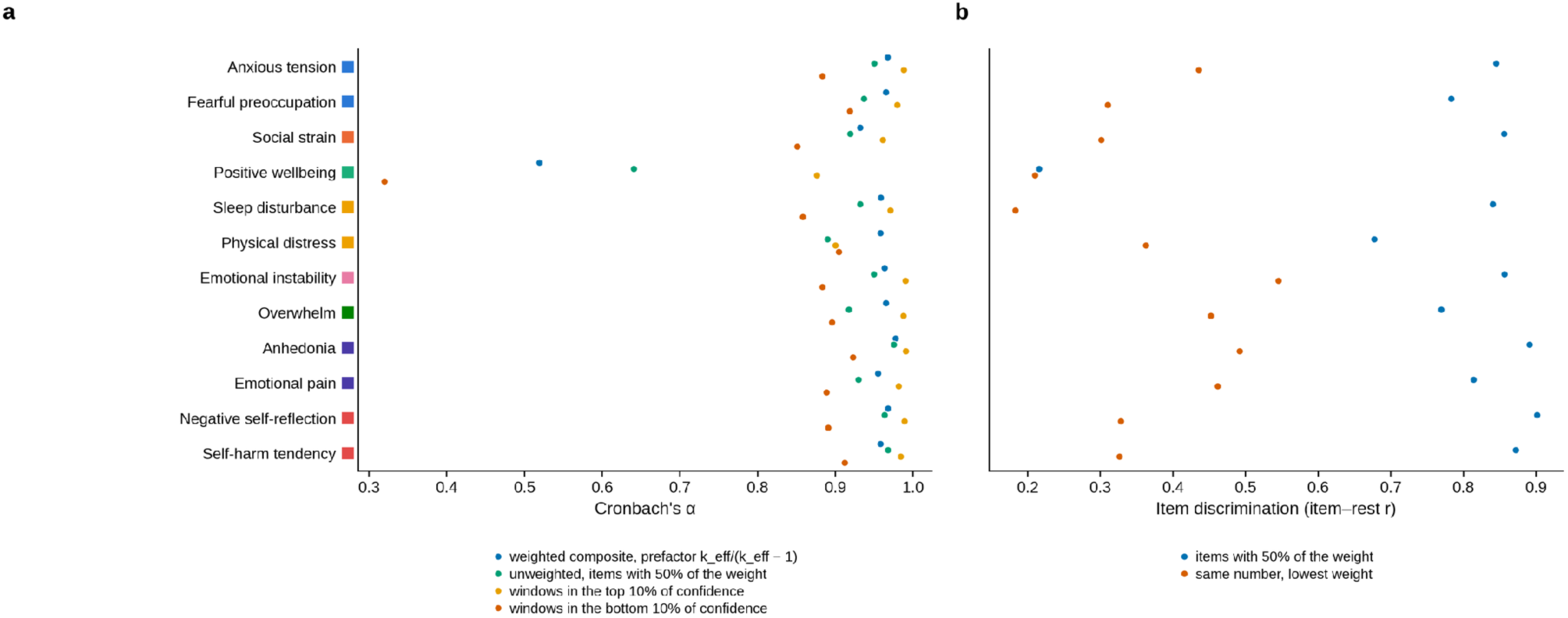
Psychometric properties under alternative estimators and confidence strata. a, Cronbach’s α computed in three ways. The main estimate is the coefficient of the weighted composite with prefactor k_eff/(k_eff − 1). The alternatives are ordinary α over the items that carry 50% of the weight and the coefficient of the weighted composite in the 10% of windows with the highest and with the lowest model confidence, weighted by the factor loadings. **b**, Discrimination of the items that carry 50% of the weight of each factor, measured by the correlation of each item with the rest of the composite, and of the same number of items with the lowest weights. Colored squares mark the clinical family of each factor as in Fig. 2.

**Extended Data Fig. 7.**
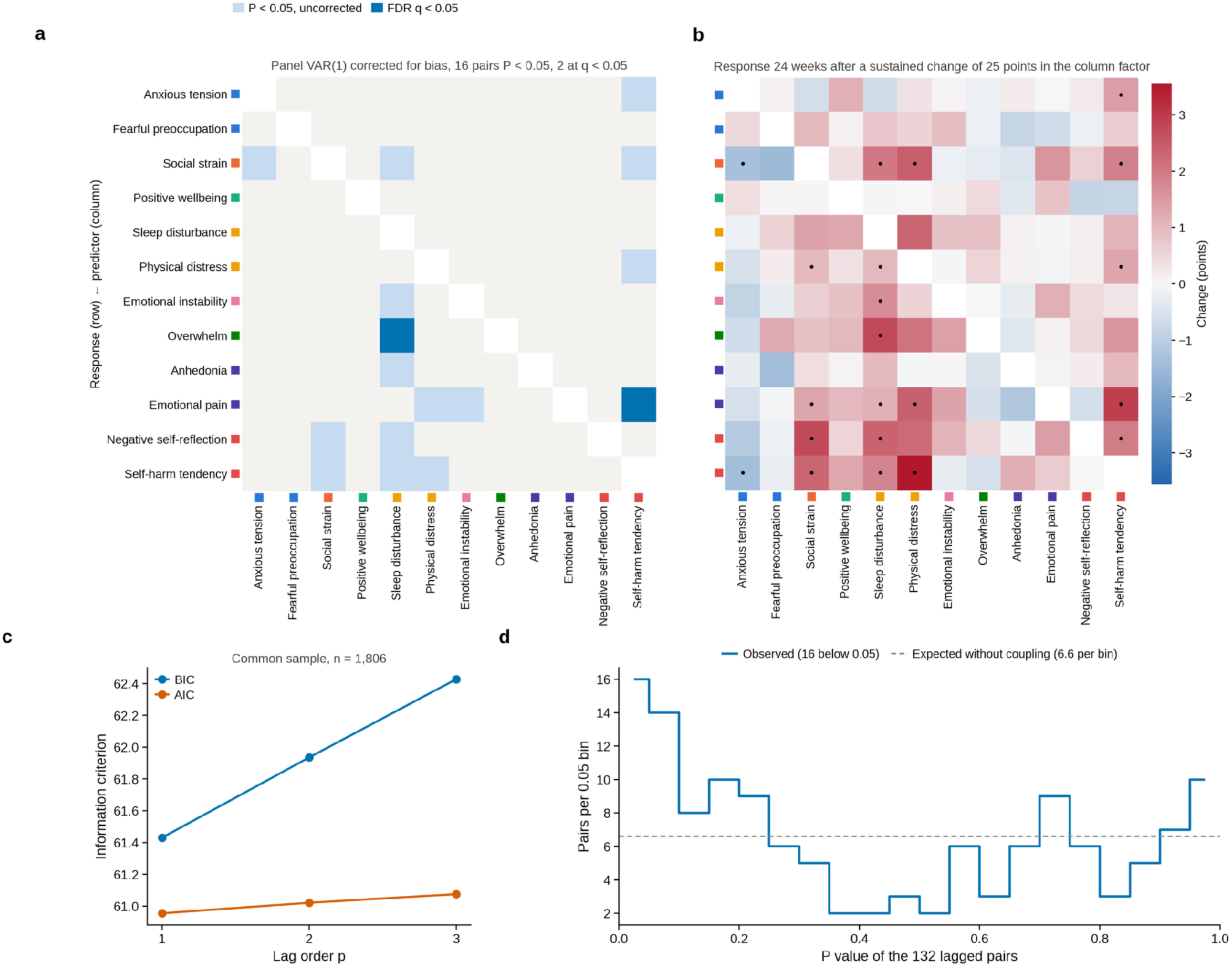
Vector autoregression within individuals. **a**, Granger predictive pairs of the panel VAR(1) with the half-panel jackknife and Wald tests based on a covariance from a bootstrap over users. Rows are the predicted factors and columns the predictors. Pale cells mark P < 0.05 without correction and dark cells FDR q < 0.05. **b**, Response of each factor (rows) 24 weeks after a sustained change of 25 points in another factor (columns). Dots mark responses whose 95% interval from a bootstrap over users excludes zero, without correction for multiple comparisons. Rows and columns are in the order of Fig. 2, and colored squares mark the clinical family of each factor. **c**, Information criteria for lag orders 1 to 3 on a common sample. **d**, Distribution of the P values of the 132 lagged pairs between factors. The dashed line marks the uniform distribution expected without any coupling.

**Extended Data Fig. 8.**
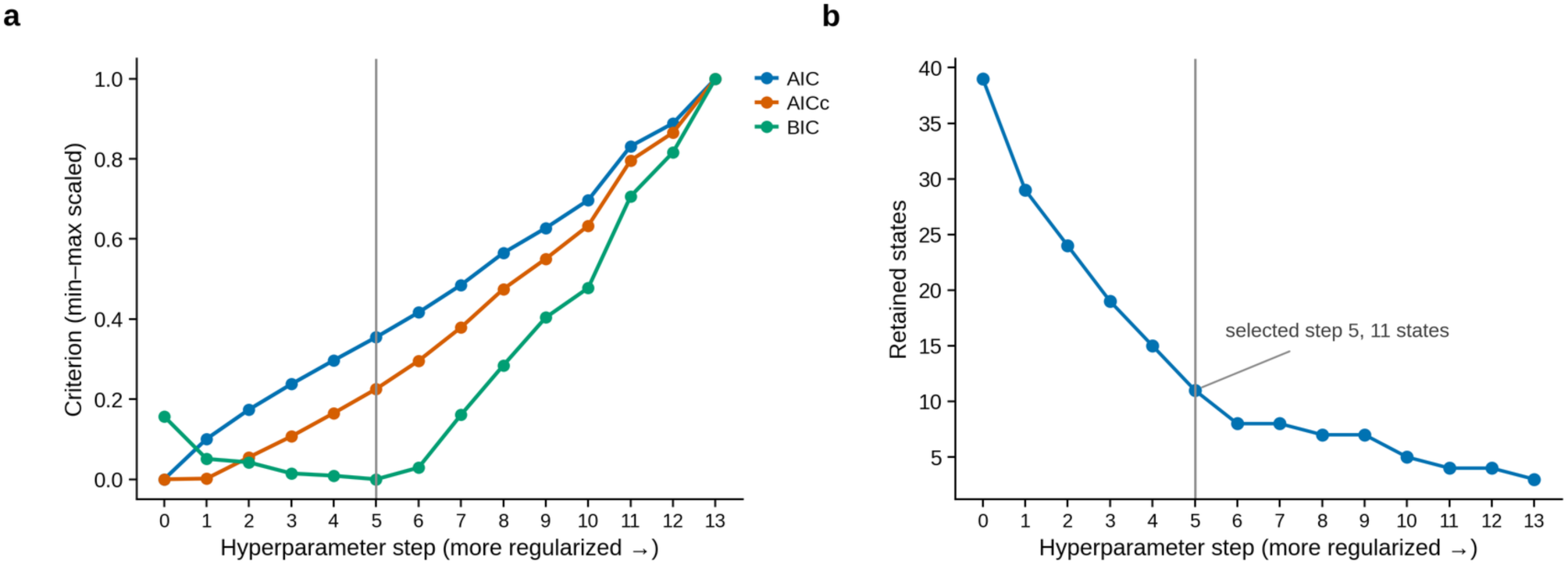
Selection of the number of discrete states. **a**, AIC, AICc and BIC across the 14 hyperparameter steps of the Dirichlet process mixture, computed with the parameter count of a Gaussian mixture with full covariances and the retained components and with the number of windows as the sample size. Each criterion is scaled to range from 0 to 1, and steps run from the least to the most regularized setting. The vertical line marks the selected step. **b**, Number of retained states at each step.

**Extended Data Fig. 9.**
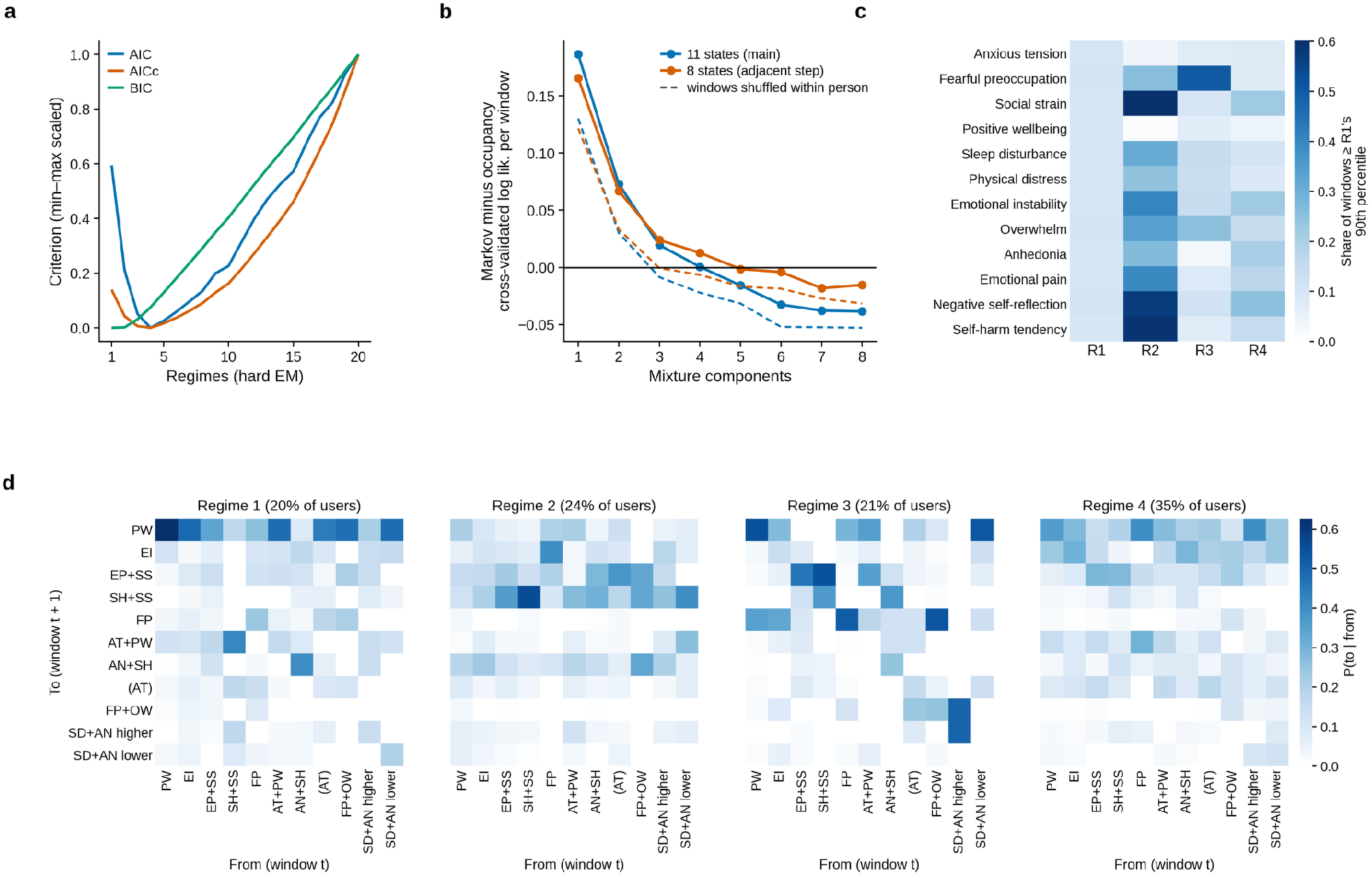
Model selection, sensitivity and description of the dynamical regimes. **a**, AIC, AICc and BIC of mixtures of Markov chains with 1 to 20 regimes, fitted with hard assignments and scaled to range from 0 to 1. **b**, Advantage in cross-validated log likelihood of Markov mixtures over occupancy mixtures by number of components, for the main solution with 11 states and for the eight states of the adjacent hyperparameter step. Dashed lines show the same after shuffling each user’s windows in time. **c**, Symptom burden, the share of each user’s windows at or above the 90th percentile of Regime 1, averaged within each regime. **d**, Transition matrices of the four regimes, P(to | from), in which a transition across missing windows counts as one step. Titles give the share of users in each regime.

**Extended Data Fig. 10.**
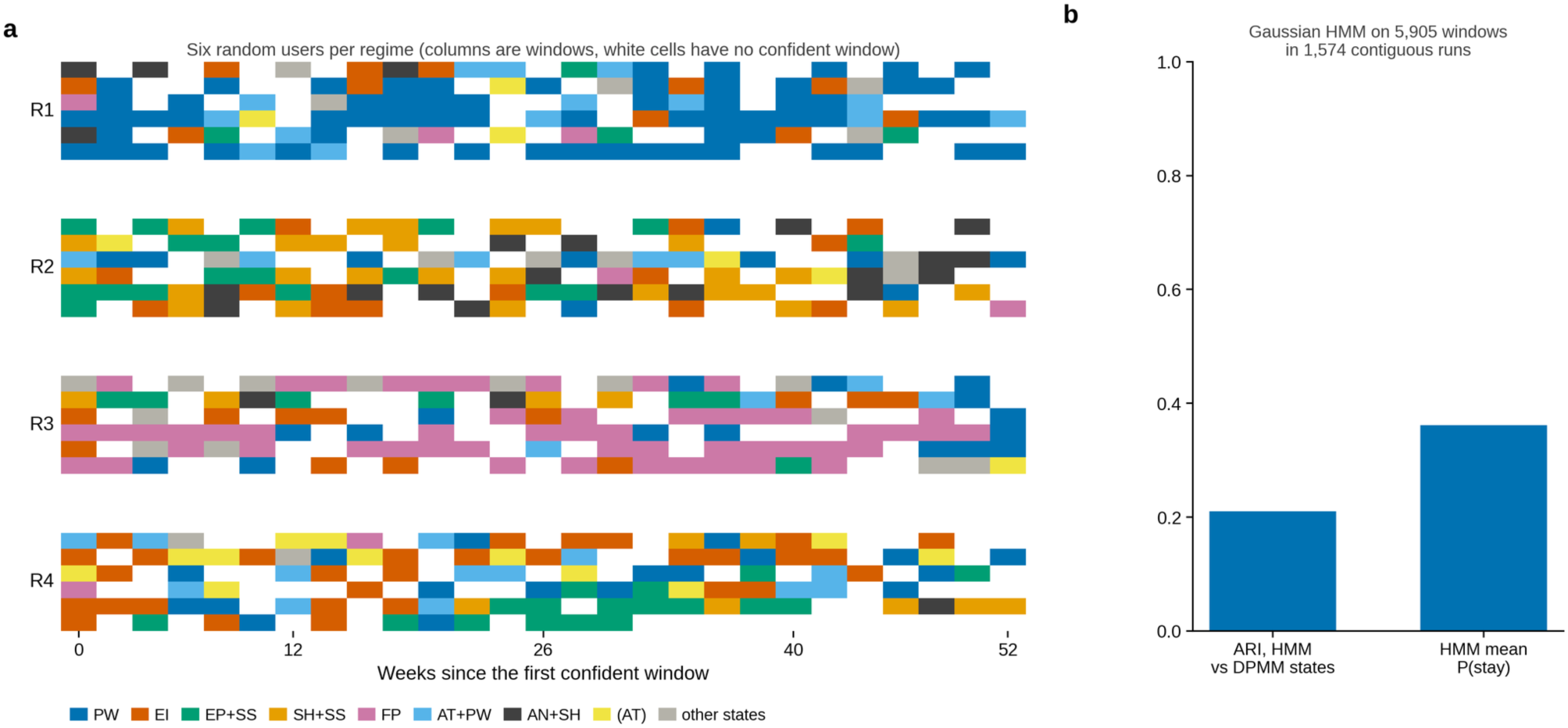
Example state sequences and a hidden Markov model. **a**, State sequences of six randomly chosen users in each regime. Columns are windows of two weeks from each user’s first confident window, and white cells mark windows without a confident rating. The eight most common states have their own colors and the remaining states are gray. **b**, A Gaussian hidden Markov model (HMM) fitted directly to the factor scores of contiguous runs of windows, with its adjusted Rand index against the states of the Dirichlet process Gaussian mixture (DPMM) and its mean probability of remaining in the same state.

## References

1. GBD 2021 Diseases and Injuries Collaborators. Global incidence, prevalence, years lived with disability (YLDs), disability-adjusted life-years (DALYs), and healthy life expectancy (HALE) for 371 diseases and injuries in 204 countries and territories and 811 subnational locations, 1990–2021: a systematic analysis for the Global Burden of Disease Study 2021. The Lancet 403, 2133–2161 (2024).

2. Cipriani, A. et al. Comparative efficacy and acceptability of 21 antidepressant drugs for the acute treatment of adults with major depressive disorder: a systematic review and network meta-analysis. The Lancet 391, 1357–1366 (2018).

3. Rush, A. J. et al. Acute and longer-term outcomes in depressed outpatients requiring one or several treatment steps: a STAR*D report. Am. J. Psychiatry 163, 1905–1917 (2006).

4. Fried, E. I., Flake, J. K. & Robinaugh, D. J. Revisiting the theoretical and methodological foundations of depression measurement. Nat. Rev. Psychol. 1, 358–368 (2022).

5. Fried, E. I. & Nesse, R. M. Depression is not a consistent syndrome: an investigation of unique symptom patterns in the STAR*D study. J. Affect. Disord. 172, 96–102 (2015).

6. Fried, E. I. The 52 symptoms of major depression: lack of content overlap among seven common depression scales. J. Affect. Disord. 208, 191–197 (2017).

7. Fried, E. I. & Nesse, R. M. Depression sum-scores don’t add up: why analyzing specific depression symptoms is essential. BMC Med. 13, 72 (2015).

8. Fortney, J. C. et al. A tipping point for measurement-based care. Psychiatr. Serv. 68, 179–188 (2017).

9. Borsboom, D. A network theory of mental disorders. World Psychiatry 16, 5–13 (2017).

10. Fried, E. I. & Robinaugh, D. J. Systems all the way down: embracing complexity in mental health research. BMC Med. 18, 205 (2020).

11. Wichers, M. The dynamic nature of depression: a new micro-level perspective of mental disorder that meets current challenges. Psychol. Med. 44, 1349–1360 (2014).

12. Nelson, B., McGorry, P. D., Wichers, M., Wigman, J. T. W. & Hartmann, J. A. Moving from static to dynamic models of the onset of mental disorder: a review. JAMA Psychiatry 74, 528–534 (2017).

13. Fisher, A. J., Medaglia, J. D. & Jeronimus, B. F. Lack of group-to-individual generalizability is a threat to human subjects research. Proc. Natl. Acad. Sci. U. S. A. 115, E6106–E6115 (2018).

14. Montgomery, S. A. & Asberg, M. A new depression scale designed to be sensitive to change. Br. J. Psychiatry J. Ment. Sci. 134, 382–389 (1979).

15. Kroenke, K., Spitzer, R. L. & Williams, J. B. W. The PHQ-9: validity of a brief depression severity measure. J. Gen. Intern. Med. 16, 606–613 (2001).

16. Beck, A. T., Steer, R. A. & Brown, G. K. Manual for the Beck Depression Inventory-II. (Psychological Corporation, 1996).

17. Insel, T. R. Digital phenotyping: technology for a new science of behavior. JAMA 318, 1215–1216 (2017).

18. Torous, J., Kiang, M. V., Lorme, J. & Onnela, J.-P. New tools for new research in psychiatry: a scalable and customizable platform to empower data driven smartphone research. JMIR Ment. Health 3, e16 (2016).

19. De Choudhury, M., Gamon, M., Counts, S. & Horvitz, E. Predicting depression via social media. Proc. Int. AAAI Conf. Web Soc. Media 7, 128–137 (2013).

20. Eichstaedt, J. C. et al. Facebook language predicts depression in medical records. Proc. Natl. Acad. Sci. U. S. A. 115, 11203–11208 (2018).

21. Chancellor, S. & De Choudhury, M. Methods in predictive techniques for mental health status on social media: a critical review. Npj Digit. Med. 3, 43 (2020).

22. Gilardi, F., Alizadeh, M. & Kubli, M. ChatGPT outperforms crowd workers for text-annotation tasks. Proc. Natl. Acad. Sci. U. S. A. 120, e2305016120 (2023).

23. Ziems, C. et al. Can large language models transform computational social science? Comput. Linguist. 50, 237–291 (2024).

24. Xu, X. et al. Mental-LLM: leveraging large language models for mental health prediction via online text data. Proc. ACM Interact. Mob. Wearable Ubiquitous Technol. 8, 31 (2024).

25. Galatzer-Levy, I. R., McDuff, D., Natarajan, V., Karthikesalingam, A. & Malgaroli, M. The capability of large language models to measure psychiatric functioning. Preprint at 10.48550/arXiv.2308.01834 (2023).

26. Obradovich, N. et al. Opportunities and risks of large language models in psychiatry. NPP—Digital Psychiatry Neurosci. 2, 8 (2024).

27. Stade, E. C. et al. Large language models could change the future of behavioral healthcare: a proposal for responsible development and evaluation. Npj Ment. Health Res. 3, 12 (2024).

28. Hamilton, M. A Rating Scale for Depression. J. Neurol. Neurosurg. Psychiatry 23, 56–62 (1960).

29. Radloff, L. S. The CES-D Scale: A Self-Report Depression Scale for Research in the General Population. Appl. Psychol. Meas. 1, 385–401 (1977).

30. Rush, A. J., Gullion, C. M., Basco, M. R., Jarrett, R. B. & Trivedi, M. H. The Inventory of Depressive Symptomatology (IDS): psychometric properties. Psychol. Med. 26, 477–486 (1996).

31. Rush, A. J. et al. The 16-Item quick inventory of depressive symptomatology (QIDS), clinician rating (QIDS-C), and self-report (QIDS-SR): a psychometric evaluation in patients with chronic major depression. Biol. Psychiatry 54, 573–583 (2003).

32. Zung, W. W. K. A Self-Rating Depression Scale. Arch. Gen. Psychiatry 12, 63 (1965).

33. Grattafiori, A., et al. The Llama 3 herd of models. Preprint at 10.48550/arXiv.2407.21783 (2024).

34. Xian, R. P. et al. Robustness tests for biomedical foundation models should tailor to specifications. Npj Digit. Med. 8, 557 (2025).

35. Garg, M., Shahbandegan, A., Chadha, A. & Mago, V. An annotated dataset for explainable interpersonal risk factors of mental disturbance in social media posts. in Findings of the Association for Computational Linguistics: ACL 2023 11960–11969 (Association for Computational Linguistics, 2023). doi:10.18653/v1/2023.findings-acl.757.

36. Garg, M., Gaur, M., Goswami, R. & Sohn, S. LoST: a mental health dataset of low self-esteem in Reddit posts. in Proceedings of the 2023 IEEE International Conference on Systems, Man, and Cybernetics 3854–3859 (IEEE, 2023). doi:10.1109/SMC53992.2023.10394671.

37. Naseem, U., Dunn, A. G., Kim, J. & Khushi, M. Early identification of depression severity levels on Reddit using ordinal classification. in Proceedings of the ACM Web Conference 2022 2563–2572 (ACM, 2022). doi:10.1145/3485447.3512128.

38. Tibshirani, R. Regression shrinkage and selection via the lasso. J. R. Stat. Soc. Ser. B Methodol. 58, 267–288 (1996).

39. Qwen Team. Qwen3.6-35B-A3B: agentic coding power, now open to all. Qwen https://qwen.ai/blog?id=qwen3.6-35b-a3b (2026).

40. Wu, S. et al. Stability-driven nonnegative matrix factorization to interpret spatial gene expression and build local gene networks. Proc. Natl. Acad. Sci. U. S. A. 113, 4290–4295 (2016).

41. Cahill, R., et al. Unsupervised pattern identification in spatial gene expression atlas reveals mouse brain regions beyond established ontology. Proc. Natl. Acad. Sci. 121, e2319804121 (2024).

42. American Psychiatric Association. Depressive disorders. in Diagnostic and Statistical Manual of Mental Disorders, Fifth Edition, Text Revision (DSM-5-TR) (American Psychiatric Association Publishing, 2022). doi:10.1176/appi.books.9780890425787.x04_Depressive_Disorders.

43. American Psychiatric Association. Anxiety disorders. in Diagnostic and Statistical Manual of Mental Disorders, Fifth Edition, Text Revision (DSM-5-TR) (American Psychiatric Association Publishing, 2022). doi:10.1176/appi.books.9780890425787.x05_Anxiety_Disorders.

44. Lorenzo-Seva, U. & ten Berge, J. M. F. Tucker’s congruence coefficient as a meaningful index of factor similarity. Methodology 2, 57–64 (2006).

45. Dhaene, G. & Jochmans, K. Split-panel jackknife estimation of fixed-effect models. Rev. Econ. Stud. 82, 991–1030 (2015).

46. Shneidman, E. S. Suicide as psychache. J. Nerv. Ment. Dis. 181, 145–147 (1993).

47. Kivelä, L. M. M., Fried, E. I., van der Does, W. & Antypa, N. Examining contemporaneous and temporal associations of real-time suicidal ideation using network analysis. Psychol. Med. 54, 3357–3365 (2024).

48. Scott, L. N. & Brown, S. L. Revisiting the affect regulation model of suicide ideation: bidirectional effects of momentary suicide ideation and affect. Suicide Life. Threat. Behav. 55, e70027 (2025).

49. Ballard, E. D. et al. Prospective association of psychological pain and hopelessness with suicidal thoughts. J. Affect. Disord. 308, 243–248 (2022).

50. Cadez, I., Heckerman, D., Meek, C., Smyth, P. & White, S. Model-based clustering and visualization of navigation patterns on a web site. Data Min. Knowl. Discov. 7, 399–424 (2003).

51. Hurvich, C. M. & Tsai, C.-L. Regression and time series model selection in small samples. Biometrika 76, 297–307 (1989).

52. Muthén, B. & Shedden, K. Finite mixture modeling with mixture outcomes using the EM algorithm. Biometrics 55, 463–469 (1999).

53. Kotov, R. et al. The Hierarchical Taxonomy of Psychopathology (HiTOP): a dimensional alternative to traditional nosologies. J. Abnorm. Psychol. 126, 454–477 (2017).

54. Van Orden, K. A. et al. The interpersonal theory of suicide. Psychol. Rev. 117, 575–600 (2010).

55. Baglioni, C. et al. Insomnia as a predictor of depression: a meta-analytic evaluation of longitudinal epidemiological studies. J. Affect. Disord. 135, 10–19 (2011).

56. Konjarski, M., Murray, G., Lee, V. V. & Jackson, M. L. Reciprocal relationships between daily sleep and mood: a systematic review of naturalistic prospective studies. Sleep Med. Rev. 42, 47–58 (2018).

57. Freeman, D. et al. The effects of improving sleep on mental health (OASIS): a randomised controlled trial with mediation analysis. Lancet Psychiatry 4, 749–758 (2017).

58. Bringmann, L. F. et al. A network approach to psychopathology: new insights into clinical longitudinal data. PLoS ONE 8, e60188 (2013).

59. Nickell, S. Biases in dynamic models with fixed effects. Econometrica 49, 1417–1426 (1981).

60. Dejonckheere, E. et al. Complex affect dynamics add limited information to the prediction of psychological well-being. *Nat*. Hum. Behav. 3, 478–491 (2019).

61. Musliner, K. L., Munk-Olsen, T., Eaton, W. W. & Zandi, P. P. Heterogeneity in long-term trajectories of depressive symptoms: patterns, predictors and outcomes. J. Affect. Disord. 192, 199–211 (2016).

62. Bauer, D. J. & Curran, P. J. Distributional assumptions of growth mixture models: implications for overextraction of latent trajectory classes. Psychol. Methods 8, 338–363 (2003).

63. Gallegos, I. O. et al. Bias and fairness in large language models: a survey. Comput. Linguist. 50, 1097–1179 (2024).

64. Xue, Z., Abbasi-Asl, R. & Pedarsani, R. Benchmarking and Mitigating the Impact of Noisy User Prompts in Medical VLMs via Cross-Modal Reflection. in Proceedings of the 19th Conference of the European Chapter of the Association for Computational Linguistics (Volume 5: Industry Track) (eds Matusevych, Y., Eryiğit, G. & Aletras, N.) 900–914 (Association for Computational Linguistics, Rabat, Morocco, 2026). doi:10.18653/v1/2026.eacl-industry.67.

65. Xue, Z., Abbasi-Asl, R. & Pedarsani, R. Enhancing the Safety of Medical Vision-Language Models by Synthetic Demonstrations. in Proceedings of the 19th Conference of the European Chapter of the Association for Computational Linguistics (Volume 1: Long Papers) (eds Demberg, V., Inui, K. & Marquez, L.) 4207–4220 (Association for Computational Linguistics, Rabat, Morocco, 2026). doi:10.18653/v1/2026.eacl-long.197.

66. Xian, R. P., et al. Assessing biomedical knowledge robustness in large language models by query-efficient sampling attacks. Trans. Mach. Learn. Res. https://openreview.net/forum?id=pvol5JyVYB (2024).

67. Chang, J. P. et al. ConvoKit: a toolkit for the analysis of conversations. in Proceedings of the 21st Annual Meeting of the Special Interest Group on Discourse and Dialogue 57–60 (Association for Computational Linguistics, 2020). doi:10.18653/v1/2020.sigdial-1.8.

68. McGraw, K. O. & Wong, S. P. Forming inferences about some intraclass correlation coefficients. Psychol. Methods 1, 30–46 (1996).

69. Lee, D. D. & Seung, H. S. Learning the parts of objects by non-negative matrix factorization. Nature 401, 788–791 (1999).

70. Pedregosa, F. et al. Scikit-learn: machine learning in Python. J. Mach. Learn. Res. 12, 2825–2830 (2011).

71. Cronbach, L. J. Coefficient alpha and the internal structure of tests. Psychometrika 16, 297–334 (1951).

72. Lütkepohl, H. New Introduction to Multiple Time Series Analysis. (Springer, 2005). doi:10.1007/978-3-540-27752-1.

73. Hamaker, E. L., Kuiper, R. M. & Grasman, R. P. P. P. A critique of the cross-lagged panel model. Psychol. Methods 20, 102–116 (2015).

74. Benjamini, Y. & Hochberg, Y. Controlling the false discovery rate: a practical and powerful approach to multiple testing. J. R. Stat. Soc. Ser. B Methodol. 57, 289–300 (1995).

75. McInnes, L., Healy, J., Saul, N. & Großberger, L. UMAP: uniform manifold approximation and projection. J. Open Source Softw. 3, 861 (2018).

76. Blei, D. M. & Jordan, M. I. Variational inference for Dirichlet process mixtures. Bayesian Anal. 1, 121–143 (2006).

77. Hubert, L. & Arabie, P. Comparing partitions. J. Classif. 2, 193–218 (1985).

78. Rabiner, L. R. A tutorial on hidden Markov models and selected applications in speech recognition. Proc. IEEE 77, 257–286 (1989).

79. Hutto, C. & Gilbert, E. VADER: a parsimonious rule-based model for sentiment analysis of social media text. Proc. Int. AAAI Conf. Web Soc. Media 8, 216–225 (2014).

80. Fast, E., Chen, B. & Bernstein, M. S. Empath: understanding topic signals in large-scale text. in Proceedings of the 2016 CHI Conference on Human Factors in Computing Systems 4647–4657 (ACM, 2016). doi:10.1145/2858036.2858535.

81. Rude, S., Gortner, E.-M. & Pennebaker, J. Language use of depressed and depression-vulnerable college students. Cogn. Emot. 18, 1121–1133 (2004).

82. Al-Mosaiwi, M. & Johnstone, T. In an absolute state: elevated use of absolutist words is a marker specific to anxiety, depression, and suicidal ideation. Clin. Psychol. Sci. 6, 529–542 (2018).

83. Seabold, S. & Perktold, J. Statsmodels: econometric and statistical modeling with Python. in Proceedings of the 9th Python in Science Conference 92–96 (SciPy, 2010). doi:10.25080/Majora-92bf1922-011.

