## Supplementary Information for "TALQ: A reproducible language model questionnaire for dense measurement of depressive and anxious states"

This file contains Supplementary Note 1 (prompts and generation settings), Supplementary Note 2 (conceptual background), Supplementary Table 1 (the 52 symptoms), Supplementary Table 2 (prespecified mapping of validation labels to symptoms), Supplementary Table 3 (prespecified mapping of inventory items to battery items) and Supplementary Table 4 (the agreement between the five ratings by the model's confidence).

### Supplementary Note 1. TALQ prompts and generation settings

**System message.** In the template below, `{posts_text}` stands for the posts and comments of the window. Each was enclosed in single quotes, and they were separated by blank lines in the order of the corpus.

```
You are playing the role of mental health professional working to help a particular client by
understanding their inner mental state. Your client has written posts on the social media site
Reddit over the last two weeks, which are shown below:

"""
{posts_text}
"""

In giving your answers about this person's inner mental state, please rely solely on evidence
that is present in the provided posts in drawing inferences about how they feel and would
respond.

Please format output in the form of a JSON object with the fields specified in the question.
```

**User message.** `{symptom_title}` and `{symptom_description}` come from Supplementary Table 1. `{symptom_scale_text}` is the rubric, which was the same for every symptom.

```
- 0: Symptom is entirely absent
- 100: Symptom is present every day
```

Typographical errors in the prompts, including the title of symptom 17 ("Indiciciveness") and "stronly" below, are reproduced as they were shown to the model.

```
You are filling out a questionnaire about your client's inner mental state over the last two
weeks, quantifying the degree to which you believe certain symptoms have been present. You are
currently thinking about the following symptom:

"""
# {symptom_title}

{symptom_description}
"""

The questionnaire instructs you to rate this symptom on a scale from 0 to 100. You can provide
any integer value in that interval as a score. The following rubric is provided in order:

"""
{symptom_scale_text}
"""

Using this scale, how would you rate your client on this symptom, given the information from
their Reddit posts over the last two weeks?

Format output in the form of a JSON object with the following fields:
* `reasoning`: (string) A four-sentence description of your internal
reasoning about what you would consider in your rating; please be brief in your response, and
limit this reasoning to a maximum of four sentences. Be sure to only have at most four sentences
in your reasoning.
```

```
* `rating`: (int) The numerical value you would select, from 0 to 100, in response to the
provided question.
* `confidence`: (int) A numerical value, from 0 to 100, indicating how stronly you believe the
given `rating` is justified from the evidence in the given Reddit posts, where, for example, a
confidence of 0 indicates that there is no supporting evidence for this judgment, 50 indicates
that there are a couple pieces of weak or indirect evidence, and 100 indicates that the Reddit
posts explicitly specify this symptom as you've rated it.
```

The system and user messages were rendered with the chat template of the model, with the generation prompt appended. Because the rendered text was then tokenized with special tokens, every input began with two tokens marking the beginning of the sequence.

**Generation settings.** The cohort annotation used Llama-3.1-8B-Instruct quantized to 4 bits in the NF4 format (bitsandbytes with transformers 4.44.2) and the default generation configuration of the checkpoint (temperature 0.6, nucleus sampling with p = 0.9, top k sampling with k = 50 and a repetition penalty of 1.0). It generated at most 384 new tokens and five samples per window and symptom. The validation on a local computer used the same weights quantized in the Q4_K_M format and served by llama.cpp, with the same sampling settings, one sample per request and a seed specific to each request. Each validation post was rated five times on each scored symptom, as in the cohort, and each calibration window once. The additional model (Qwen3.6-35B-A3B) used weights quantized to about 4 bits and served by llama.cpp on the same computer, its own chat template with its thinking mode switched off, the same sampling settings and a seed specific to each request, and it rated each post once.

### Supplementary Note 2. Conceptual background on language models as fixed interpreters of mental life

This note sets out the conceptual motivation for treating a language model as a fixed rater of mental states. It also discusses the clinical interpretation of two of the factors.

**Language as a record of shared mental structure.** How can the experiences of different people be compared when subjective experience is accessible only to the person who has it? Language has long been central to this question. One proposal is that languages, like other cultural artifacts, carry an image of the relational structure of human mental life. A model trained to reproduce the statistics of language might therefore encode some of that structure.

Whether large language models understand language in any substantive sense is actively debated.[1] Our results do not settle that debate, and they do not need to, because what matters for measurement is narrower. The judgments of the model were aggregated over repeated samples[2] and anchored to everything one person wrote over two weeks, a median of 417 words. The resulting ratings were reliable and agreed with constructs annotated by humans in independent posts. Their factor structure was close to established symptom taxonomies and departed from them in ways that can be interpreted.

**A fixed interpreter.** Every measure of a mental state relates a person's inner life to a reference concept, such as negative self-reflection, and both self-report and clinician ratings pass through an interpreter. In self-report, the interpreter is the person, whose reading of each item depends on their history and current state. In clinician ratings, the interpreter is the clinician, whose reading depends on training and on inferential biases.

A language model is also an interpreter, with biases of its own, and its semantics differ from those of any person. Unlike a human rater, however, it can be fixed. Its weights can be archived, its idiosyncrasies characterized and its judgments repeated exactly years later. Variability between raters is replaced by a single source of bias that can be inspected. That bias can then be studied, for example by validation against human labels (Fig. 1d) and by comparing models.

**Two refinements of canonical symptomatology.** The factor analysis separated Anxious tension (interpersonal sensitivity, feeling bothered and inner tension) from Fearful preoccupation (illness anxiety, panic and sympathetic arousal). The latter plausibly indexes anxious states of high arousal,[3] and sympathetic activation has been implicated in illness anxiety.[4] Inner tension, by contrast, has been treated as a basic dimension of feeling since the beginnings of experimental psychology,[5] and recent accounts link it to uncertainty about future states.[6]

Social strain captured negative judgments attributed to others ("other people dislike me"), as distinct from negative judgments of oneself. Sociology has long distinguished the self as both judge and judged from the self as judged by another.[7] This distinction is absent from the criteria for major depression, and social anxiety disorder concerns fear of social situations rather than the inferred hostility of others. An index of perceived social evaluation may therefore have research and clinical value of its own.

**From states to dynamics.** The proposal of dynamical regimes moves the definition of a phenotype from particular states to the way states unfold over time. Epilepsy offers a model for this view.[8] A single seizure does not define epilepsy. The disorder is an altered propensity to enter seizure states, and the field has moved from one concept to many epilepsies, distinguished by their patterns of transition and their semiology. At the resolution of two weeks in the present data, however, regimes added little to state occupancy (Fig. 5), so the proposal remains to be tested with denser measurement.

The same logic applies to behavioral neuroscience. Segmentation of animal behavior into discrete states and their transitions[9,10] has so far concentrated on time scales from below a second to minutes. TALQ extends this approach to time scales of weeks to months, at which comparable slow phenotypes could also be sought in model organisms.

### Supplementary Table 1. The 52 symptoms rated by TALQ

Descriptions are those shown to the model, including their typographical errors. Titles are also as shown, except that the title of symptom 17 is given in its correct spelling here and was shown to the model as "Indiciciveness". The 12 symptoms marked 'title only' were presented by their title alone. Every item used the same rubric with two anchors (Supplementary Note 1). Where figures use a shorter name, it is given in parentheses.

| No. | Symptom (name in figures) | Description shown to the model |
|---|---|---|
| 1 | Initial insomnia | Difficulty falling asleep at the beginning of the night |
| 2 | Sleep maintenance insomnia (Middle insomnia) | Difficulty maintaining sleep in the middle of the night; waking up multiple times in the middle of the night |
| 3 | Late insomnia | Difficulty with early morning awakenings; waking up before one intended to, and being unable to fall back asleep |
| 4 | Hypersomnia | Excessive daytime sleepiness despite getting adequate sleep; falling asleep multiple times during the day |
| 5 | Sad mood | *(title only)* |
| 6 | Anxiousness / Fearful mood (Anxious mood) | Apprehension or uneasiness that stems from the anticipation of danger, which may be internal or external |
| 7 | Panic / Phobic symptoms (Panic / phobic) | Sense of immediately impending doom or danger; fear of loss of control or death; shortness of breath or throat tightness; dizziness, lightheadedness, ro faintness. Can be accompanied by feelings of unreality or detachment. |
| 8 | Irritable mood | Proneness to anger from small frustrations or negative events |
| 9 | Emotional reactivity | Responding immediately with strong emotions to small events or other stimuli, with decreased ability to regulate this emotional response |
| 10 | Diurnal variation | *(title only)* |
| 11 | Grief | Subjective feeling that emotional responses are tied to a personal loss and ensuing feelings of absence |
| 12 | Decreased appetite | *(title only)* |
| 13 | Increased appetite | *(title only)* |
| 14 | Weight loss | *(title only)* |
| 15 | Weight gain | *(title only)* |
| 16 | Concentration problems | Difficulty in focusing on the task at hand while ignoring distractions, especially for longer periods or more intense tasks |
| 17 | Indecisiveness | Inability to, or difficulty in, making any kind of choice in the client's daily life |
| 18 | Guilt | Client feels that they are emotionally responsible for negative outcomes in their own life or the lives of others around them, even when that may not be supported by evidence or believed by other observers |
| 19 | Worthlessness | Client feels low self-esteem, that they are of little or no value as a person |
| 20 | Pessimism | Hopelessness about the future |
| 21 | Suicidal ideation | Thoughts, ideas, or ruminations that the client would be better off dead, the possibility of ending the client's own life, or specific circumstances or planned actions around ending the client's own life |

| No. | Symptom (name in figures) | Description shown to the model |
|---|---|---|
| 22 | Loss of interest | The client no longer has a desire to do things that once brought them pleasure |
| 23 | Loss of pleasure | Things that the client once experienced as pleasurable are no longer experienced as pleasurable |
| 24 | Fatigue | Persistent exhaustion, malaise, aversion to activity, or impaired performance on activities |
| 25 | Loss of energy | Loss or decreased frequency of periods of incresed drive to activity |
| 26 | Decreased libido | *(title only)* |
| 27 | Psychomotor retardation | Slowing down or reduction of thought, speech, affect, or physical movements |
| 28 | Psychomotor agitation | Excessive and uncontrolled mental and/or motor activity |
| 29 | Somatic complaints | Reports of symptoms in the body, like pain, headaches, or tingling, that are not overtly explained by another medical cause |
| 30 | Sympathetic arousal | Including palpitations, tinnitus, chest pain, tremors |
| 31 | Gastrointestinal problems (GI problems) | Including constipation, diarrhea |
| 32 | Interpersonal sensitivity | Vulnerability and increased response to perceived signs of social rejection |
| 33 | Leaden paralysis | Feeling of increased weight in the extremities that makes it more difficult to do activities |
| 34 | Emphasis on past failure (Past failure) | Persistent rumination on thoughts or memories that the client perceives as demonstrating their personal failure in the past |
| 35 | Punishment feelings | Feeling that the client is deserving of being puniushed, either in response to past failures or because of feeling intrinsically worthless |
| 36 | Self-dislike | *(title only)* |
| 37 | Self-criticalness | Persistent emphasis or rumination on how events or stimuli reflect negative aspects of the client |
| 38 | Crying | Crying more than the client has in the past, or crying at moments in which there is no stimulus or minimal stimulus evoking the response |
| 39 | Loneliness | Unpleasant emotional response or psychological pain in response to perceived social isolation |
| 40 | Excessive effort | Subjective feeling that everything the client does takes effort |
| 41 | Talking less | Client perceives that they talk less than they have in the past |
| 42 | Feeling that "Other people were unfriendly to me" (Others unfriendly) | Subjective feeling that others have acted recently in a way that was unfriendly to the client |
| 43 | Feeling that "Other people dislike me" (Others dislike me) | Subjective feeling that others dislike the client |
| 44 | Feeling bothered | Subjective feeling that things which usually wouldn't bother the client, or have not in the past, seem more bothersome at present |
| 45 | Feeling as good as other people (Feeling as good as others) | Client believes that how they feel is about as good as how the others around them in the world feel |
| 46 | Feeling happy | *(title only)* |
| 47 | Feeling useful to or needed by others (Feeling needed) | *(title only)* |

| No. | Symptom (name in figures) | Description shown to the model |
| --- | --- | --- |
| 48 | Feeling that life is full (Life overfull) | In a negative, overwhelming sense: that is, the client reports that their life is over-full of too many different things |
| 49 | Inner tension | Feelings of ill-defined discomfort, inner turmoil, edginess, mounting panic or dread |
| 50 | Inability to feel | Client does not have emotional responses to stimuli around them, positive or negative; experiencing emotional "numbness" |
| 51 | Hypochondriasis | *(title only)* |
| 52 | Loss of insight | *(title only)* |

### Supplementary Table 2. Prespecified mapping of validation labels to TALQ symptoms and factors

The mapping, including the control symptoms, was specified before any TALQ output on the validation posts existed, and the SHA-256 hash of the mapping file is recorded in the code repository. The mapping determined which symptoms were scored on each dataset, whereas the predictive models of Fig. 1d combined all scored symptoms without using it. Each composite is the mean of the listed symptoms, and inverse items enter as 100 minus the rating. The control composite is the mean of three symptoms without a conceptual link to the labels and was scored for the datasets on interpersonal risk factors and low self-esteem. The mapping file also specified labels of two further datasets that were not scored in this study.

| Dataset | Label | Symptoms (inverse items marked $^{-}$) | Factor | Composite |
|---|---|---|---|---|
| Interpersonal risk factors[11] | Thwarted belongingness | Loneliness, Feeling that "Other people dislike me", Feeling that "Other people were unfriendly to me", Interpersonal sensitivity | Social strain | Mean |
| Interpersonal risk factors[11] | Perceived burdensomeness | Worthlessness, Punishment feelings, Guilt, Suicidal ideation, Feeling useful to or needed by others $^{-}$ | Negative self-reflection | Mean |
| LoST, low self-esteem[12] | Low self-esteem | Worthlessness, Self-dislike, Self-criticalness, Emphasis on past failure, Feeling as good as other people $^{-}$ | Negative self-reflection | Mean |
| Depression severity[13] | Severity (minimum, mild, moderate, severe) | Sad mood, Loss of interest, Loss of pleasure, Decreased appetite, Increased appetite, Weight loss, Weight gain, Initial insomnia, Sleep maintenance insomnia, Late insomnia, Hypersomnia, Psychomotor agitation, Psychomotor retardation, Fatigue, Loss of energy, Worthlessness, Guilt, Concentration problems, Indecisiveness, Suicidal ideation | None | Mean |
| Interpersonal risk factors and LoST | Control composite | Initial insomnia, Decreased appetite, Gastrointestinal problems | None | Mean |

### Supplementary Table 3. Prespecified correspondence between inventory items and TALQ battery items

The correspondence was written from the titles and wording of the items before any inventory rating was compared with the battery, and the SHA-256 hash of the mapping file is recorded in the code repository. Where an inventory item covers several battery items, they are combined by their mean, or by their maximum for items that cover changes in either direction (sleep, appetite and psychomotor activity).

| Inventory | Item | Title | TALQ battery items | Combination |
|---|---|---|---|---|
| BDI-II | 1 | Sadness | Sad mood | Single item |
| BDI-II | 2 | Pessimism | Pessimism | Single item |
| BDI-II | 3 | Past Failure | Emphasis on past failure | Single item |
| BDI-II | 4 | Loss of Pleasure | Loss of pleasure | Single item |
| BDI-II | 5 | Guilty Feelings | Guilt | Single item |
| BDI-II | 6 | Punishment Feelings | Punishment feelings | Single item |
| BDI-II | 7 | Self-Dislike | Self-dislike | Single item |
| BDI-II | 8 | Self-Criticalness | Self-criticalness | Single item |
| BDI-II | 9 | Suicidal Thoughts or Wishes | Suicidal ideation | Single item |
| BDI-II | 10 | Crying | Crying | Single item |
| BDI-II | 11 | Agitation | Psychomotor agitation | Single item |
| BDI-II | 12 | Loss of Interest | Loss of interest | Single item |
| BDI-II | 13 | Indecisiveness | Indecisiveness | Single item |
| BDI-II | 14 | Worthlessness | Worthlessness | Single item |
| BDI-II | 15 | Loss of Energy | Loss of energy | Single item |
| BDI-II | 16 | Changes in Sleeping Pattern | Initial insomnia, Sleep maintenance insomnia, Late insomnia, Hypersomnia | Maximum |
| BDI-II | 17 | Irritability | Irritable mood | Single item |
| BDI-II | 18 | Changes in Appetite | Decreased appetite, Increased appetite | Maximum |
| BDI-II | 19 | Concentration Difficulty | Concentration problems | Single item |
| BDI-II | 20 | Tiredness or Fatigue | Fatigue | Single item |
| BDI-II | 21 | Loss of Interest in Sex | Decreased libido | Single item |
| PHQ-9 | 1 | Anhedonia | Loss of interest, Loss of pleasure | Mean |
| PHQ-9 | 2 | Depressed mood | Sad mood, Pessimism | Mean |
| PHQ-9 | 3 | Sleep disruption | Initial insomnia, Sleep maintenance insomnia, Late insomnia, Hypersomnia | Maximum |
| PHQ-9 | 4 | Fatigue | Fatigue, Loss of energy | Mean |
| PHQ-9 | 5 | Appetite disruption | Decreased appetite, Increased appetite | Maximum |
| PHQ-9 | 6 | Worthlessness / guilt | Worthlessness, Guilt, Emphasis on past failure | Mean |

| Inventory | Item | Title | TALQ battery items | Combination |
|---|---|---|---|---|
| PHQ-9 | 7 | Concentration | Concentration problems | Single item |
| PHQ-9 | 8 | Psychomotor retardation | Psychomotor retardation, Psychomotor agitation | Maximum |
| PHQ-9 | 9 | Suicidality | Suicidal ideation | Single item |

### Supplementary Table 4. Agreement between the five ratings by the model's confidence.

Each window–symptom pair of the cohort with five valid ratings (507,994 pairs) was assigned to a group by the model's mean confidence over the five ratings, on a scale of 0 to 100. Groups include their lower bound and exclude their upper bound. Share is the percentage of pairs in each group. Agreement is the ICC(1) of single ratings, computed for each symptom with at least 30 pairs in the group and summarized as the median over symptoms. The 86 pairs with confidence below 30 were too few to estimate agreement and are not shown. The main analyses kept windows in which the mean confidence exceeded 40 for every symptom.

| Model confidence | Share of ratings | Agreement of the five ratings (median ICC) |
|---|---|---|
| 30–40 | 0.5% | 0.19 |
| 40–50 | 2.8% | 0.29 |
| 50–60 | 9.1% | 0.37 |
| 60–70 | 26% | 0.48 |
| 70–80 | 34% | 0.73 |
| 80–100 | 28% | 0.90 |

#### References.


1. Mitchell, M. & Krakauer, D. C. The debate over understanding in AI's large language models. Proc. Natl Acad. Sci. USA 120, e2215907120 (2023).

2. Farquhar, S., Kossen, J., Kuhn, L. & Gal, Y. Detecting hallucinations in large language models using semantic entropy. Nature 630, 625–630 (2024).

3. Kreibig, S. D. Autonomic nervous system activity in emotion: a review. Biol. Psychol. 84, 394–421 (2010).

4. Abramowitz, J. S., Schwartz, S. A. & Whiteside, S. P. A contemporary conceptual model of hypochondriasis. Mayo Clin. Proc. 77, 1323–1330 (2002).

5. Wundt, W. Grundriss der Psychologie (Engelmann, 1896).

6. Lehne, M. & Koelsch, S. Toward a general psychological model of tension and suspense. Front. Psychol. 6, 79 (2015).

7. Goffman, E. The Presentation of Self in Everyday Life (Doubleday, 1959).

8. Engel, J. Jr. A proposed diagnostic scheme for people with epileptic seizures and with epilepsy: report of the ILAE Task Force on Classification and Terminology. Epilepsia 42, 796–803 (2001).

9. Wiltschko, A. B. et al. Mapping sub-second structure in mouse behavior. Neuron 88, 1121–1135 (2015).

10. Ashwood, Z. C. et al. Mice alternate between discrete strategies during perceptual decision-making. Nat. Neurosci. 25, 201–212 (2022).

11. Garg, M., Shahbandegan, A., Chadha, A. & Mago, V. An annotated dataset for explainable interpersonal risk factors of mental disturbance in social media posts. In Findings of the Association for Computational Linguistics: ACL 2023 11960–11969 (ACL, 2023).

12. Garg, M., Gaur, M., Goswami, R. & Sohn, S. LoST: a mental health dataset of low self-esteem in Reddit posts. In Proc. 2023 IEEE International Conference on Systems, Man, and Cybernetics 3854–3859 (IEEE, 2023).

13. Naseem, U., Dunn, A. G., Kim, J. & Khushi, M. Early identification of depression severity levels on Reddit using ordinal classification. In Proc. ACM Web Conference 2022 2563–2572 (ACM, 2022).
